# The impact of local vaccine coverage and recent incidence on measles transmission in France between 2009 and 2018

**DOI:** 10.1101/2021.05.31.21257977

**Authors:** Alexis Robert, Adam J. Kucharski, Sebastian Funk

## Abstract

**Background:** Despite high levels of vaccine coverage, sub-national heterogeneity in immunity to measles can create pockets of susceptibility, which are hard to detect and may result in long-lasting outbreaks. The elimination status defined by the World Health Organization aims to identify countries where the virus is no longer circulating and can be verified after 36 months of interrupted transmission. However, since 2018, numerous countries have lost their elimination status soon after reaching it, showing that the indicators used to define elimination may not be predictive of lower risks of outbreaks.

**Methods and Findings:** We quantified the impact of local vaccine coverage and recent levels of incidence on the dynamics of measles in each French department between 2009 and 2018, using mathematical models based on the ‘Epidemic-Endemic’ regression framework. High values of local vaccine coverage were associated with fewer imported cases and lower risks of local transmissions. Regions that had recently reported high levels of incidence were also at a lower risk of local transmission, potentially due to additional immunity accumulated during these recent outbreaks. Therefore, all else being equal, the risk of local transmission was not lower in areas fulfilling the elimination criteria (i.e., low recent incidence). After fitting the models using daily case counts, we used the parameters’ estimates to simulate the effect of variations in the vaccine coverage and recent incidence on future transmission. A decrease of 3% in the three-year average vaccine uptake led to a five-fold increase in the number of cases simulated in a year on average.

**Conclusions:** Spatiotemporal variation in vaccine coverage because of disruption of routine immunisation programmes, or lower trust in vaccines, can lead to large increases in both local and cross regional transmission. The association found between local vaccine coverage and incidence suggests that, although regional vaccine uptake can be hard to collect and unreliable because of population movements, it can provide insights into the risks of imminent outbreak. Periods of low local measles incidence were not indicative of a decrease in the risks of local transmission. Therefore, the incidence indicator used to define the elimination status was not consistently associated with lower risks of measles outbreak in France. More detailed models of local immunity levels or subnational seroprevalence studies may yield better estimates of local risk of measles outbreaks.

## Introduction

Immunity against infectious diseases accumulates following infection and, if a vaccine is available, routine immunisation programs and vaccination campaigns. Measles is highly infectious and can cause large outbreaks in populations with low immunity [1,2]. Therefore, high levels of vaccine coverage are required to minimise the risks of outbreaks [3]. Furthermore, vaccine uptake must be homogeneously high across the territory to avoid local transmission sustained by regional discrepancies [4,5]. The large-scale implementation of routine immunisation programs led to a drastic reduction in measles cases worldwide, and measles was targeted for elimination in five World Health Organization (WHO) Regions by 2020 under the Global Vaccine Action Plan 2011-2020 [6].

Elimination status, as defined by the WHO, refers to “the absence of endemic measles transmission for ≥12 months in the presence of a well-performing surveillance system” in a given country or region, and is verified “after 36 months of interrupted endemic measles virus transmission”[7]. Although imported cases, or cases directly related to importations could still be expected, there should be no continuous transmission persisting over a long period of time in a region where measles was eliminated. A given WHO region can declare measles eliminated when all countries in the region document interruption of endemic transmission for more than 36 months.

Recently, several countries had their elimination status revoked following large outbreaks less than five years after it was verified. For instance, the United Kingdom achieved elimination in 2017, and lost the status in 2019 along with Albania, Czechia, Greece, Venezuela, and Brazil [8,9]. In these countries, interruption of transmission during a few years was not indicative of reduced risks of major outbreaks. Such occurrences can be explained by several factors, such as a replenishment of susceptible individuals after years without transmission, or importations of cases into subnational areas with lower levels of immunity caused by heterogeneity in vaccine coverage [10–13]. The number and geographical distribution of the susceptible individuals is not routinely monitored in most countries given the perceived cost and logistical challenges of large serological surveys, yet it is a main predictor of outbreak risk [3]. Local values of vaccine coverage can be an alternative measure of heterogeneity, but they are not always available and can be outdated because of the mobility between regions. Furthermore, they only describe vaccine-induced immunity, and therefore ignore the immunity caused by previous outbreaks. In this study, we aim to i) estimate the impact of recent local transmission and local vaccine coverage on the current risk of outbreaks, and the changes in transmission dynamics that would results from variations in these factors, and ii) identify the areas most at-risk for local transmission using France as a case study.

To do so, we implemented an Epidemic-Endemic time-series model using *hhh4*, a framework developed by Held, Höhle and Hofmann to study the separate impact of covariates on importation, cross-regional transmission and local transmissions on aggregated case counts [14,15]. We adapted this framework to daily case counts and applied it to the daily number of measles cases per department (NUTS3 levels) in France reported to the European Center for Disease Prevention and Control (ECDC) between January 2009 and December 2018. We computed the average values of vaccine uptake and the number of cases per department in the past three years to mimic the timeframe used to define the elimination status, and modelled their impact on the local risks of outbreaks.

## Methods

### Description of the hhh4 framework

We used the modelling framework implemented in the “hhh4” model, which is part of the R package “*surveillance*”[15], to analyse infectious disease case counts. All the notations are defined in Table 1. The expected number of cases (*μ*_*i,t*_) reported in the region *i* at time *t* depends on three sources of transmission (called “components”):

**Table 1:**
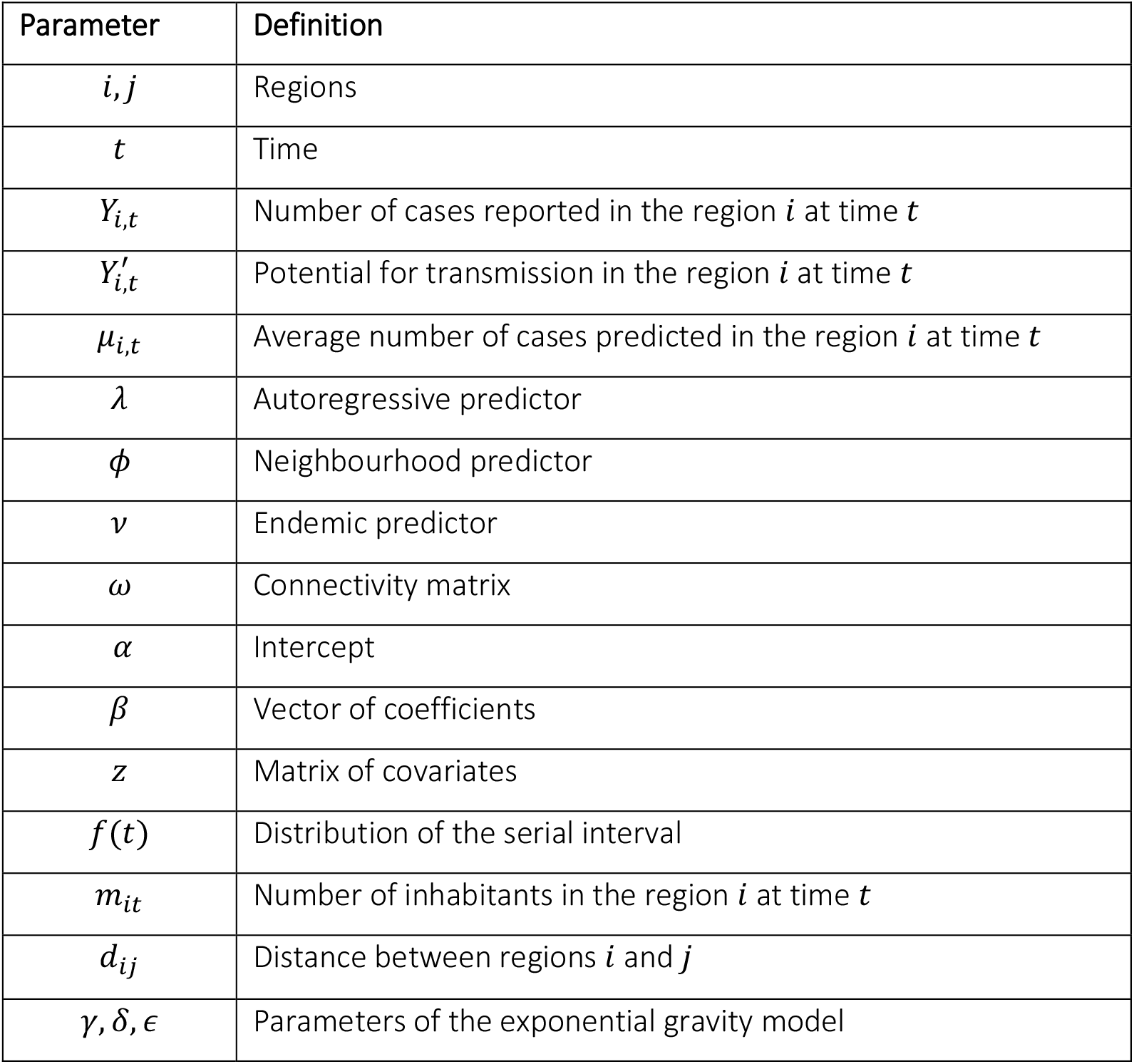
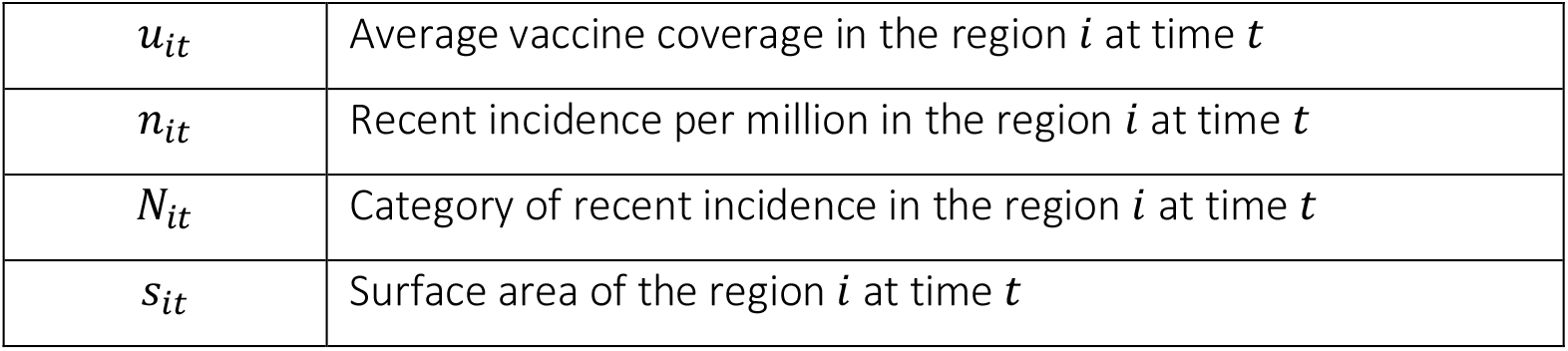
Table of notations of all variables and distributions defined in the methods.

| Parameter | Definition |
| --- | --- |
| $i, j$ | Regions |
| $t$ | Time |
| $Y_{i,t}$ | Number of cases reported in the region $i$ at time $t$ |
| $Y'_{i,t}$ | Potential for transmission in the region $i$ at time $t$ |
| $\mu_{i,t}$ | Average number of cases predicted in the region $i$ at time $t$ |
| $\lambda$ | Autoregressive predictor |
| $\phi$ | Neighbourhood predictor |
| $v$ | Endemic predictor |
| $\omega$ | Connectivity matrix |
| $\alpha$ | Intercept |
| $\beta$ | Vector of coefficients |
| $z$ | Matrix of covariates |
| $f(t)$ | Distribution of the serial interval |
| $m_{it}$ | Number of inhabitants in the region $i$ at time $t$ |
| $d_{ij}$ | Distance between regions $i$ and $j$ |
| $\gamma, \delta, \epsilon$ | Parameters of the exponential gravity model |
| $u_{it}$ | Average vaccine coverage in the region $i$ at time $t$ |
| $n_{it}$ | Recent incidence per million in the region $i$ at time $t$ |
| $N_{it}$ | Category of recent incidence in the region $i$ at time $t$ |
| $s_{it}$ | Surface area of the region $i$ at time $t$ |

i. The *autoregressive* component (*λ*_*i,t*_) represents the impact of *Y*_*i,t*−1_, the number of cases in *i* at the previous time step, on the number of cases in *i* at *t*. The number of new cases expected from the autoregressive component is the product of predictors *λ*_*i,t*_ and *Y*_*i,t*−1_. A high value of *λ*_*i,t*_ indicates that, if there are cases in *i*, there is potential for high transmission levels. On the other hand, if *λ*_*i,t*_ is low, cases in *i* are unlikely to lead to much local transmission.
ii. The *neighbourhood* component (*ϕ*_*i,t*_) represents the impact of *Y*_*j,t*−1_, the number of cases reported in regions around *i* at the previous time step, on the number of cases in *i* at *t*. The exact impact of cases in these regions on cases in *i* is determined by a distance matrix *ω* which quantifies the connectivity between the different regions. If *ϕ*_*i,t*_ is high, cases in regions around *i* are more likely to cause new cases in *i*, whereas a low value of *ϕ*_*i,t*_ indicates that cross regional transmissions towards *i* are less likely.
iii. The *endemic* component (*v*_*i,t*_) represents the background number of new cases occurring in region *i*, regardless of the current number of cases in *i*, or in the regions around *i*. If *v*_*i,t*_ is high, new cases in *i* are common, regardless of the number of cases in or around *i* at the previous time step. Since the endemic component does not depend on *Y*_*t*−1_, it represents the background importations that cannot be linked to the mechanistic components. Therefore, these cases either correspond to importations from outside the modelled area (France in our case), or cases that are not otherwise predicted by the other two components.

The full equation for the expected number of cases in region *i* at time *t* is:

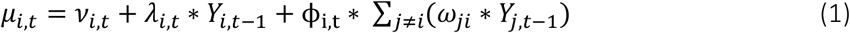

The predictors *λ*_*i,t*_, *ϕ*_*i,t*_ and *v*_*i,t*_ are independently impacted by different covariates, i.e., a covariate may be associated with a reduction of importations, but have little impact on the spread of the virus within the region. We assume that *Y*_*i,t*_, the number of observed cases at *t* in *i*, follows a negative binomial distribution to allow for overdispersion [16]. The overdispersion parameter *ψ* is estimated.

The predictors *λ*_*i,t*_, *ϕ*_*i,t*_ and *v*_*i,t*_ are estimated using log-linear regressions. For each predictor, we estimate: i) The intercept *α* (identical across spatial units), and ii) the vector of coefficients *β* associated with *z*_*i,t*_ the vector of covariates at *t* in *i* included in each component.

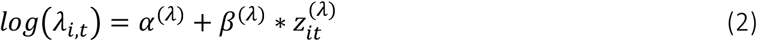

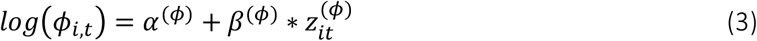

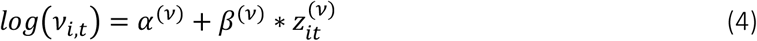

### Data

The observed case counts *Y*_*i,t*_ was computed from 14,461 cases (10,988 confirmed and 3,473 probable cases) routinely collected in metropolitan France, and reported to the ECDC between January 2009 and December 2018 (Figure 1A). This data was retrieved on The European Surveillance System (TESSy) on 22 January 2019. The cases were stratified by the metropolitan department they were reported in. The department correspond to French NUTS3 regions. We excluded three cases where this information was not available. We used the date of symptom onset reported for each case to compute the daily number of cases from 2009 to 2018 per department.

**Figure 1:**
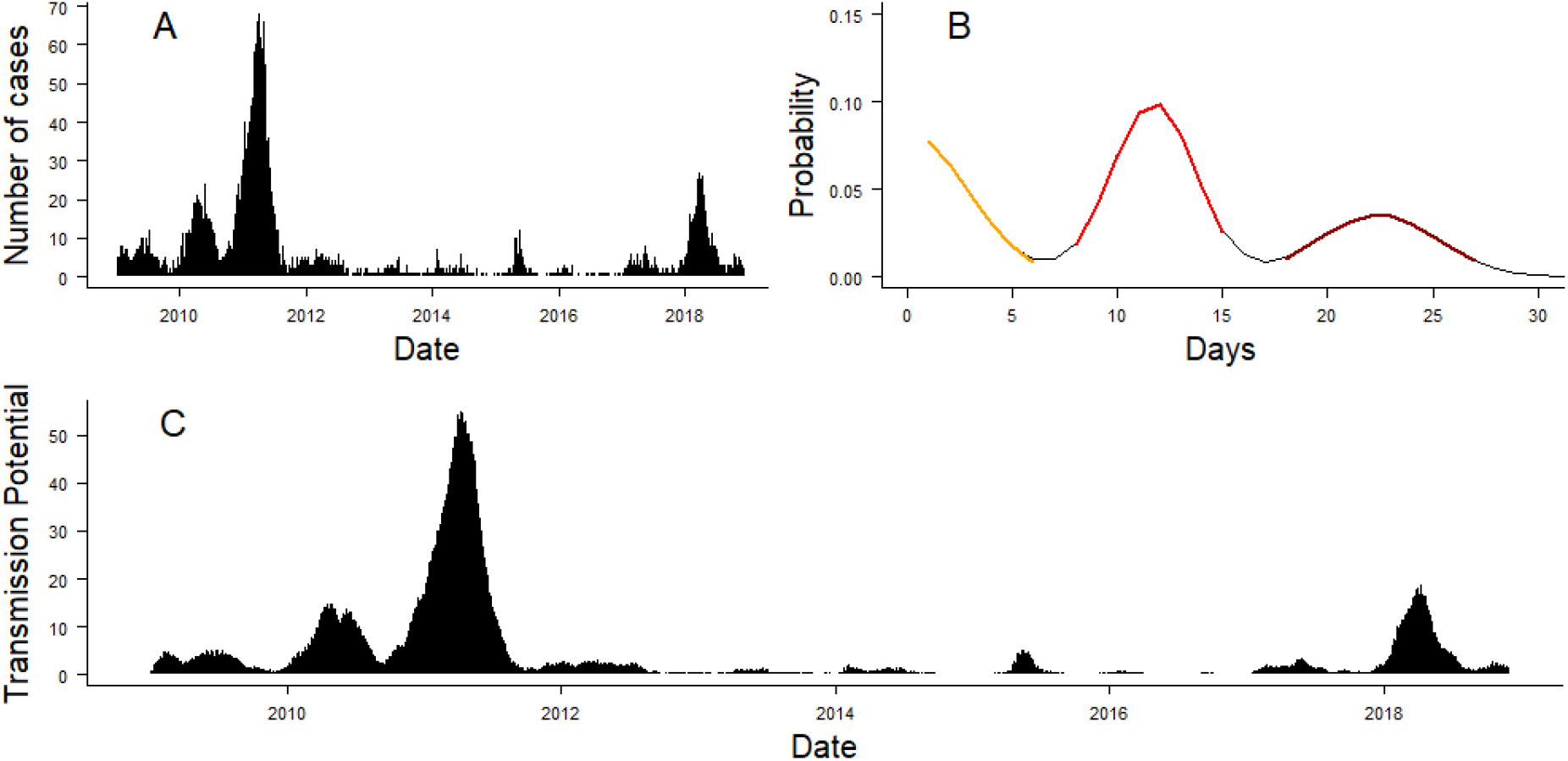
Panel A: Daily number of cases reported in France between 1^st^ January 2009 and 30^th^ November 2018. Panel B: Distribution of the composite serial interval used in the model. The different colours of the curve correspond to the three scenarios used to compute the distribution of the serial interval (orange: serial interval when missing ancestor; red: serial interval without unreported case, brown: serial interval when the case between the two reported cases was missing). Panel C: Transmission potential, which was computed by convolving the number of cases in the last 30 days with the composite serial interval.

### Adaptation of hhh4 to daily case counts

In *hhh4*, the average number of new cases stemming from the autoregressive and neighbourhood components depends on the number of cases at the previous time step. Therefore, if we use daily case counts, the number of cases at *t* is only impacted by the number of cases the day before. In reality, however, the serial interval of measles is estimated to be 11 days on average [17]. Previous studies using *hhh*4 relied on temporally aggregated case counts, which partially solved this problem: if the time step is close to the average serial interval, cases of the same generation of transmission can be assumed to be roughly grouped together in the same time point [18]. Nevertheless, studying weekly (or fortnightly) aggregated cases counts does not reflect the distribution of the serial interval (i.e., it ignores overlapping generations of transmission because of shorter or longer delays between primary and secondary cases). This can lead to directly connected cases being grouped in the same time step, or separated by more than one time step. This aggregation also ignores the potential for unreported cases, which may lead to cases causing transmission two to three weeks after their onset date via an intermediate, unobserved case. Finally, the starting date of aggregation influences how cases are grouped, which can lead to discrepancies in the parameter estimates.

Recent developments in the *surveillance* package included weight estimation to represent the relative impact of previous time steps on the number of cases at *t* [19]. Since we are using daily case counts, we set the weights of the different time steps from the distribution of the serial interval. We computed *Y*′_*it*_, the transmission potential for each department and time step, by multiplying the number of recent cases by the distribution of the serial interval 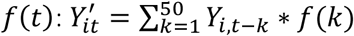 Only a subset of measles cases are reported to the surveillance system [20], therefore we accounted for the risks of unreported cases by computing a composite serial interval from three different transmission scenarios (Figure 1B):

1. In case of direct transmission between two cases *i* and *j*, the number of days between the two cases *f*_1_(*t*) follows a Normal distribution truncated at 0: *f*_1_(*t*)∼*N*(11.7, 2) [17].
2. In case of unreported cases between *i* and *j*, the number of days between the two cases *f*_2_(*t*) follows a Normal distribution truncated at 0: *f*_2_(*t*)∼*N*(23.4, √8). This distribution corresponds to the convolution of *f*_1_(*t*) with itself.
3. If *i* and *j* share the same unreported index case, the number of days between *i* and *j* follows a half-Normal distribution (excluding 0) of standard deviation √8 days. This distribution corresponds to the distribution of the difference of *f*_1_(*t*) with itself, excluding values below 1. We added this last scenario to account for multiple concurrent importations stemming from an unreported infector.

We considered that 50% of the composite serial interval reflected direct transmission (scenario 1, without missing generations between cases), and 50% came from the two scenarios with unreported cases (scenarios 2 and 3). The distribution of the composite serial interval is shown in Figure 1B. We ran sensitivity analysis to estimate the parameters of the model using composite serial intervals computed with different proportions of direct transmission, and observed it had little influence on the estimation of each parameter (Supplement Section 1).

### Connectivity between departments

In the *hhh4* framework, the average number of cases caused in the department *i* at time *t* by cases from another department *j* is quantified by the neighbourhood component. It is equal to ϕ_i,t_ ∗ *ω*_*ji*_ ∗ *Y*_*j,t*−1_ (Equation 1). Therefore, the number of cases caused by cases from *j* in *i* in *hhh4* is influenced by three factors:

- The susceptibility of the department *i*, quantified by the neighbourhood predictor ϕ_i,t_, defined as 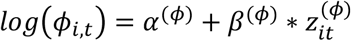.
- The number of connections from *j* to *i*, calculated using an exponential gravity model [21], whereby the number of connections between *i* and *j* is proportional to the product of the number of inhabitants in the department of origin *m*_*j*_, the department of destination *m*_*i*_ and an exponential decrease in the distance between *i* and *j d*_*ji*_. Therefore, the number of connections from *j* to *i* was calculated as 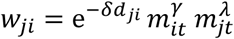.
- The proportion of the population in *j* that is infectious.

Therefore, the average number of cases expected from department *j* to department *i* at *t* can be written as the product of these three factors:

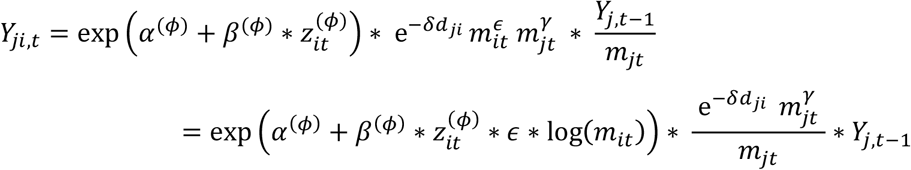

Therefore, the log-population log (*m*_*it*_) was added as a covariate of the predictor of the neighbourhood component *ϕ*. The number of inhabitants in each French department between 2009 and 2018 was taken from the INSEE website [22].

We implemented two models with different methods to compute the distance between departments *d*_*ji*_.

1. In Model 1, every department can be connected to each other, therefore only importations coming from outside the departments included in the study fall into the endemic component. The distance matrix was computed using the distance between the population centroids of each department, which were calculated using the 1*km*^2^ European Grid dataset [23]. This dataset contains the number of inhabitants in each grid cell covering the country (resolution 1km). We computed the weighted population centre in each department using the R function *zonal* from the package raster[24] and calculated the distance between population centres.

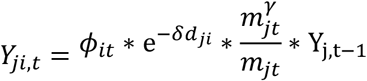
2. In Model 2, the neighbourhood component only takes into account transmission between neighbouring departments, assuming that cross-regional transmissions between non-neighbouring departments would be captured by the baseline number of daily importations (i.e. the endemic component):

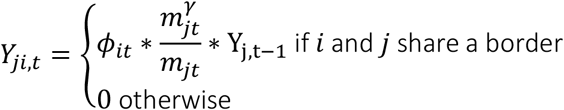

Therefore, the neighbourhood component in Model 1 includes both the neighbourhood component and part of the endemic transmission in Model 2.

### Covariates

Different covariates can be added in each component of the *hhh4* framework [25]. We implemented the same set of covariates in the two models. The two covariates of interest were the impact of vaccine coverage and the category of incidence in each department in the past three years. We chose this timeframe in order to match the requirements of the elimination status assessment. We also included the number of inhabitants, the surface area of each department, and the seasonality as control variables, as explained below:

#### Vaccine coverage

For each department *i* and time step *t*, we computed *u*_*i,t*_, the average proportion unvaccinated in the department *i* over the 3 years prior to *t* according to local coverage reports. We averaged over the past three years in order to use the same timeframe as the elimination status assessment. We used the yearly first dose uptake among 2-year-old children in each French department between 2006 and 2017. This data is publicly available on the website Santé Publique France [26–28]. The uptake of the second dose was not reported before 2010, and many departments had missing entries after 2010. Therefore, only the local coverage of the first dose was used in the model.

Since 26% of the entries in the coverage dataset were missing, we ran a beta mixed model to infer the missing values. We used the time and squared time (in years) as covariates, and random effects stratified by department. We used the average prediction to infer the missing values from the fitted model and get the complete vaccine coverage dataset. More details on the regression, and the sensitivity analyses that were run are presented in the Appendix (Supplement Section 2). All values of coverage in 2009 were missing, and were not imputed; we computed the average vaccine coverage in 2010, 2011, and 2012 using only two of the three previous years.

Adding the log-proportion of unvaccinated to the model was the most appropriate approach, since it allows the rate of disease spread (i.e. the value of the predictors *λ, v*, and *ϕ*) to be proportional to the density of susceptibles [25]. Therefore, we calculated the average log-proportion of unvaccinated in the three years before *t* and added it as a covariate in all three components.

#### Impact of recent incidence

This covariate quantifies the impact of past outbreaks on current transmission. Departments are eligible for WHO certification of elimination status if they have maintained low levels of transmission over the past three years [7]. Therefore, we computed *n*_*i,t*_, the number of cases per million reported between a month and three years before *t* in *i*. We excluded cases reported in the last month since recent cases may be directly linked to current transmission.

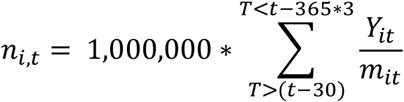

We aggregated n_it_ in three categories: i)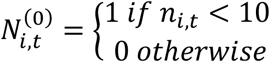 :very limited transmission in recent years, department potentially eligible for elimination (30% of entries) ; ii) 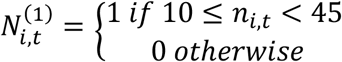 : Moderate transmission in recent years (36% of entries); iii) 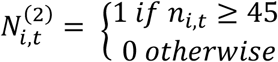 : major outbreak reported in the department in recent years. The threshold of 45 cases per million corresponds to the last tercile of *n*_*i,t*_, hence 33% of *n*_*i,t*_ fall into this last category.

Computing the level of recent incidence required the number of cases per department in the past three years. Therefore, since this analysis integrates case counts data from 2009, we needed to compute the incidence in each department between 2006 and 2008. Less than 50 cases were reported in France per year in 2006 and 2007 [29], therefore we considered their contribution to the recent level of incidence per department was null. On the other hand, 597 measles cases were reported to the ECDC in France in 2008, but were not stratified by department. Therefore, we used the number of cases reported per department in 2008 on Sante-Publique-France (597 cases overall, mostly reported in the second half of 2008 [30]) and integrated them in the computation of *N*_*i,t*_ for *t* < 2012.

The level of recent incidence was a covariate in all three components.

#### Number of inhabitants and surface area

In the subsection “Connectivity between departments”, we discussed the impact of the number of inhabitants on the number of movements between departments. Furthermore, several studies have indicated a potential association between the population density and the number of secondary transmissions [31–33]. Therefore, we controlled for the impact of the number of inhabitants in each department, and the surface area (i.e., the geographical size) on the number of local transmissions.

The log-number of inhabitants log (*m*_*i,t*_) in the department *i* at time *t* was added as a covariate in all three components. The log-surface of the department log (*s*_*i,t*_) was added as a covariate in the autoregressive component.

#### Seasonality

We control for the impact of the seasonality of measles outbreaks in France on transmission by adding two covariates (sine-cosine) to all three components.

#### Full model equations for predictors

The covariates are all integrated in the covariate vectors in the equations 2, 3 and 4, yielding:

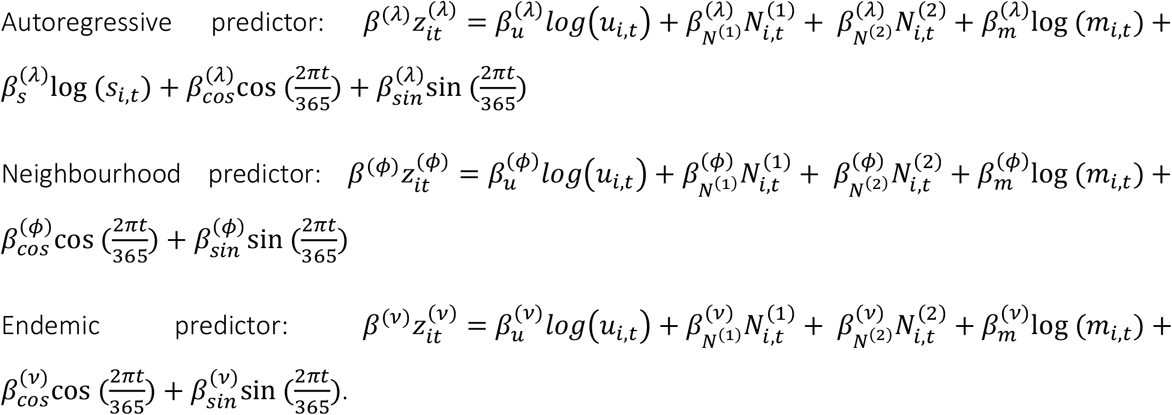

### Model calibration

A model is deemed well-calibrated if it is able to correctly identify its own uncertainty in making predictions [34]. The most straightforward method to evaluate whether *hhh*4 models are well-calibrated is to generate a one-step-ahead forecast over a chosen test period and compare them with the data [15]. Since we use daily case counts, this method would only assess the ability of the models to capture the number of cases on the next day. We explored the calibration of our models several days ahead. To do so, we selected the last two years of data as the test period, fit the model up to each day, and simulated the number of cases over the next 3, 7, 10 and 14 days for each day of the test period in each department. For each date, we ran at least 100,000 simulations. If the number of cases observed in the data had not been generated in 100,000 simulations, we ran simulations until it was reached. From these simulations, we generated the predictive probability distribution at each time step in each department. In a model with perfect calibration, the actual number of cases follows the predictive probability distribution (*μ*_*it*_ ∼*P*_*it*_ for all predictive distributions *P*_*it*_), i.e., the probability integral transform (PIT) histogram is uniform. We computed the PIT histograms in both models for predictions over 3, 7, 10, and 14 days. The PIT histograms were computed using a non-randomised yet uniform version of the PIT histogram correcting for the use of discrete values described in Czado et al [35] and implemented in *hhh4*.

The PIT histograms were used to estimate whether the short-term forecasts were in line with the data, and whether the models were consistently missing some scenarios of transmission.

### Simulation study

In order to highlight the impact of variations in the local vaccine coverage or the level of recent transmission on the risks of outbreaks, we generated simulations of the number of cases in France across one year under different conditions. To compute these simulations, we used the last values of average vaccine coverage (the average was computed from the values in 2015, 2016, and 2017) and the levels of recent incidence in mid-2018, and simulated the daily number of cases between the 1^st^ of August 2018 and the 31^st^ of December 2019. We started the simulations during the period of the year associated with the lowest number of cases (i.e., on the 1^st^ of August), in order to avoid biases. Indeed, if we had used the last three months of data (until November 2018), some departments may have been repeatedly associated with higher numbers of cases in our simulations, not because they are more at risk of importation or transmission, but because there had been cases reported in these departments at the beginning of the epidemic year. We were only interested in highlighting the impact of variations in coverage and recent transmission, rather than predicting the level of transmission for the entire year of 2019.

We generated 100 samples of the regression coefficients using the variance-covariance matrix and assumed they followed a multivariate normal distribution. For each sample, we computed the values of the three predictors between the 1^st^ of August 2018 and the 31^st^ of December 2019, and simulated the daily number of cases in each department across the year. We ran 100 simulations per sample (i.e. 10,000 simulations were generated per scenario).

We studied four scenarios: i) Using the latest local values of coverage (averaged over the past three years), population and category of recent incidence, ii) Increasing the vaccination coverage in each department by three percent, iii) Decreasing the vaccination coverage in each department by three percent, and iv) setting the recent incidence in each department to minimal levels (i.e. conditions fulfilling the WHO elimination status requirements).

Finally, since tourism and local events can lead to mass gatherings and trigger repeated importations independent of parameters included in the model [36,37], we studied the impact of repeated local importations of cases into specific departments. To do so, we simulated one year of transmission (i.e., until the end of 2019) following the importations of 10 cases in a given department in December 2018. In these simulations, we did not allow for any other baseline importations throughout the year, in order to assess the potential for geographical spread throughout the country after importation in one department.

## Results

### Impact of the covariates on each component

The parameter estimates obtained in both models are shown in Figure 2. Values above 0 show aggravating effects associated with an increase in the number of expected cases at the next time step. For both models, departments with a high proportion unvaccinated in the past three years were associated with a higher number of expected cases in the autoregressive (Model 1: 0.14 [0.03 - 0.24] ; Model 2: 0.19 [0.09 - 0.29]) and the endemic component (Model 1: 0.37 [-0.17 - 0.91] ; Model 2: 0.48 [0.17 - 0.80]). This indicates that these departments were at higher risks of background importations, and secondary transmission upon importation. In both components, the effect of vaccination was slightly stronger in Model 2, where cross-regional transmission is restricted to neighbouring departments, than in Model 1, where cross-regional transmission can happen between all departments, although the confidence intervals overlapped. In Model 1, the proportion unvaccinated also had an aggravating effect on the number of cross-departmental transmissions (0.47 [0.23 - 0.71]), whereas in Model 2 there was no clear association between the proportion unvaccinated and an increase in cross-regional transmission (−0.02 [-0.29 - 0.25]). The differences between the models’ coefficients were due to the cross-regional transmission in Model 1 corresponding to both the neighbourhood component and some of the endemic transmission in Model 2.

**Figure 2:**
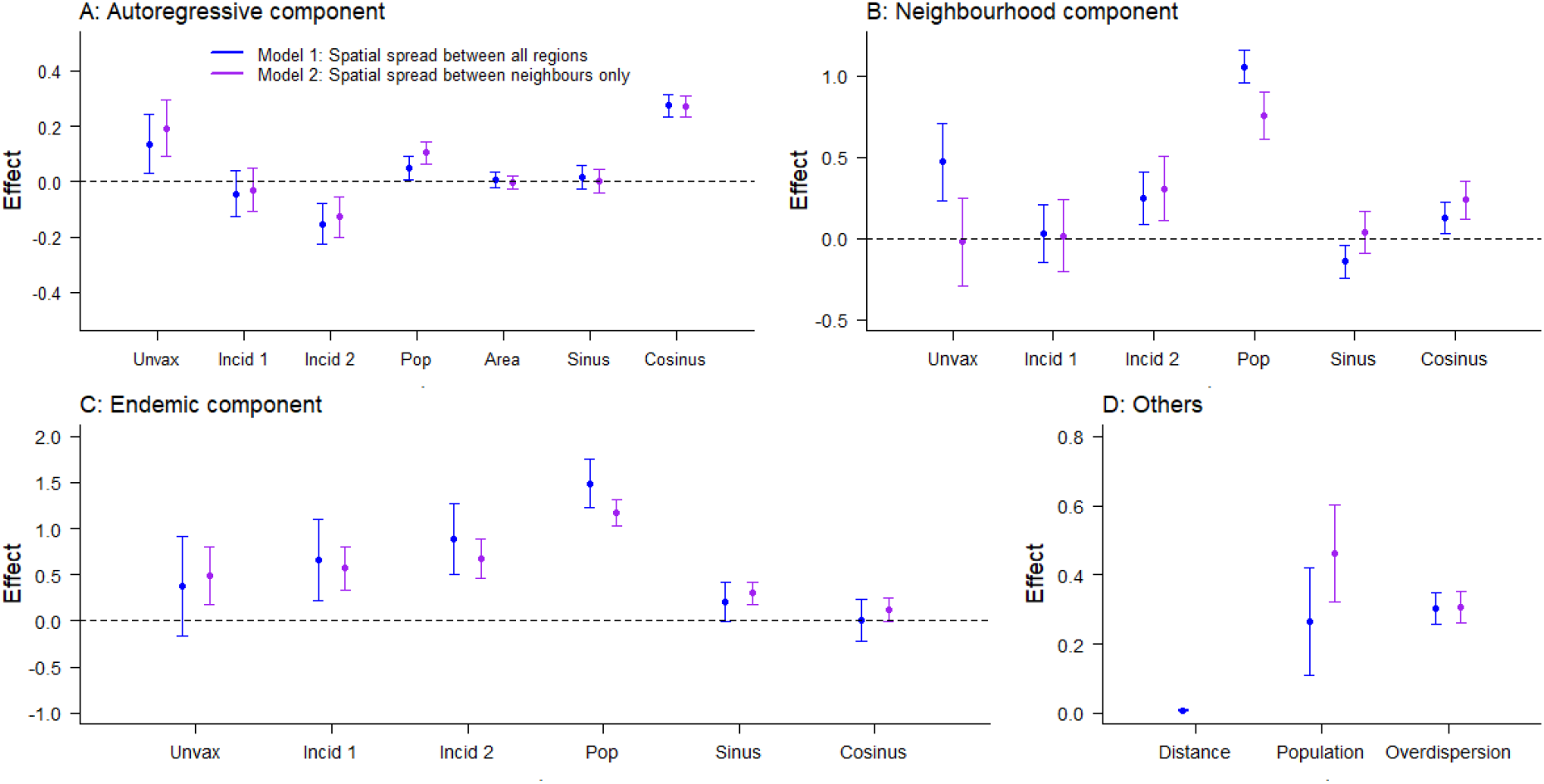
Estimates of the parameters in each component of Model 1 (blue) and Model 2 (purple): Panel A: Autoregressive component; Panel B: Neighbourhood component; Panel C: Endemic component; Panel D: Other coefficients. The y-axis. unvax corresponds to the effect of u_i,t_, the mean proportion unvaccinated over the three years before t in i; incid1 and incid2 correspond to the effect of 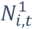 and 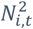 the category of incidence in the three years before t in i; pop corresponds to the effect of m_i,t_ the number of inhabitants at t in i; area corresponds to the effect of the surface; sin and cos correspond to the effects of seasonality; distance and population correspond to the spatial parameters of the connectivity matrix w (δ and γ); overdisp is the estimate of the log-overdispersion parameter in the negative binomial distribution of Y_i,t_. Dots show the mean values associated with the parameters; arrows show the 95% Confidence interval. Note different y-axes between graphs.

The association between the level of incidence over the past three years (parameters: *immun* 1 and *immun* 2 in Figure 2) and the components of transmission was similar in both models. In the auto-regressive component, departments that reported high incidence over the past three years (*immun* 2) were associated with fewer secondary cases per case in the department (Model 1: -0.15 [-0.23 - -0.08]; Model 2: -0.13 [-0.20 - -0.06]). This could be linked to outbreak-induced immunity causing a depletion of susceptibles in departments where incidence was high over the past few years. On the other hand, the parameters associated with *immun* 2 were above 0 in the neighbourhood and endemic components, which indicates that departments with high incidence in the past three years were more at risk of cross-regional transmission and background importations (Model 1: Endemic 0.89 [0.50 – 1.27]; Neighbourhood: 0.25 [0.09 – 0.41]; Model 2: Endemic 0.67 [0.46 – 0.89]; Neighbourhood: 0.31 [0.11 – 0.51]). The parameter *immun* 1 was only significantly different from 0 in the endemic component (Model 1: 0.66 [0.22 – 1.10]; Model 2: 0.57 [0.34 – 0.80]), meaning departments that recently reported moderate levels of transmission were associated with more background importations, but no difference was noticeable in cross-regional or within-region transmission.

The other covariates included in the model showed that the number of inhabitants in a department had an important impact on both the endemic and neighbourhood components: departments with more individuals were more likely to report background importations and cross-regional transmission. On the other hand, the population and the surface area of the departments had no impact on the autoregressive component. We also observed a strong impact of seasonality on the three components (Figure 2). Indeed, the peak values of the predictors were 20 to 37% higher than the average value in all components of transmission (Supplement Section 3). The peak of the autoregressive component was in February for both models, the endemic peak was in May for Model 1 (April in Model 2), whereas the neighbourhood component peaked in December in Model 1 (March in Model 2).

Using the mean parameter estimates, and the latest values of vaccination coverage, incidence, and number of inhabitants per department, we computed the local predictors *ϕ*_*i*_, *λ*_*i*_, and *v*_*i*_ in both models to highlight the spatial heterogeneity of the transmission risks (Figure 3). The predictors were computed ignoring the impact of seasonality, which does not change the geographic distribution of risks since it is not region-dependent in the models. Therefore, the maps correspond to the average local value of the predictors the year following the last data entry (i.e. the 30^th^ of November 2018). The geographic distributions of the autoregressive predictor are similar in Model 1 and Model 2. This indicates that the same departments were classified as having higher risks of local transmission in both models. Areas with lower values of vaccine uptake such as the South East and South West of France were associated with higher risks of secondary transmission. Indeed, the highest values of within-region transmission were reported in Bouches-du-Rhône and Var (in the South East of France). Populous departments in the North of France were also at risk of secondary transmission despite higher vaccination coverage.

**Figure 3:**
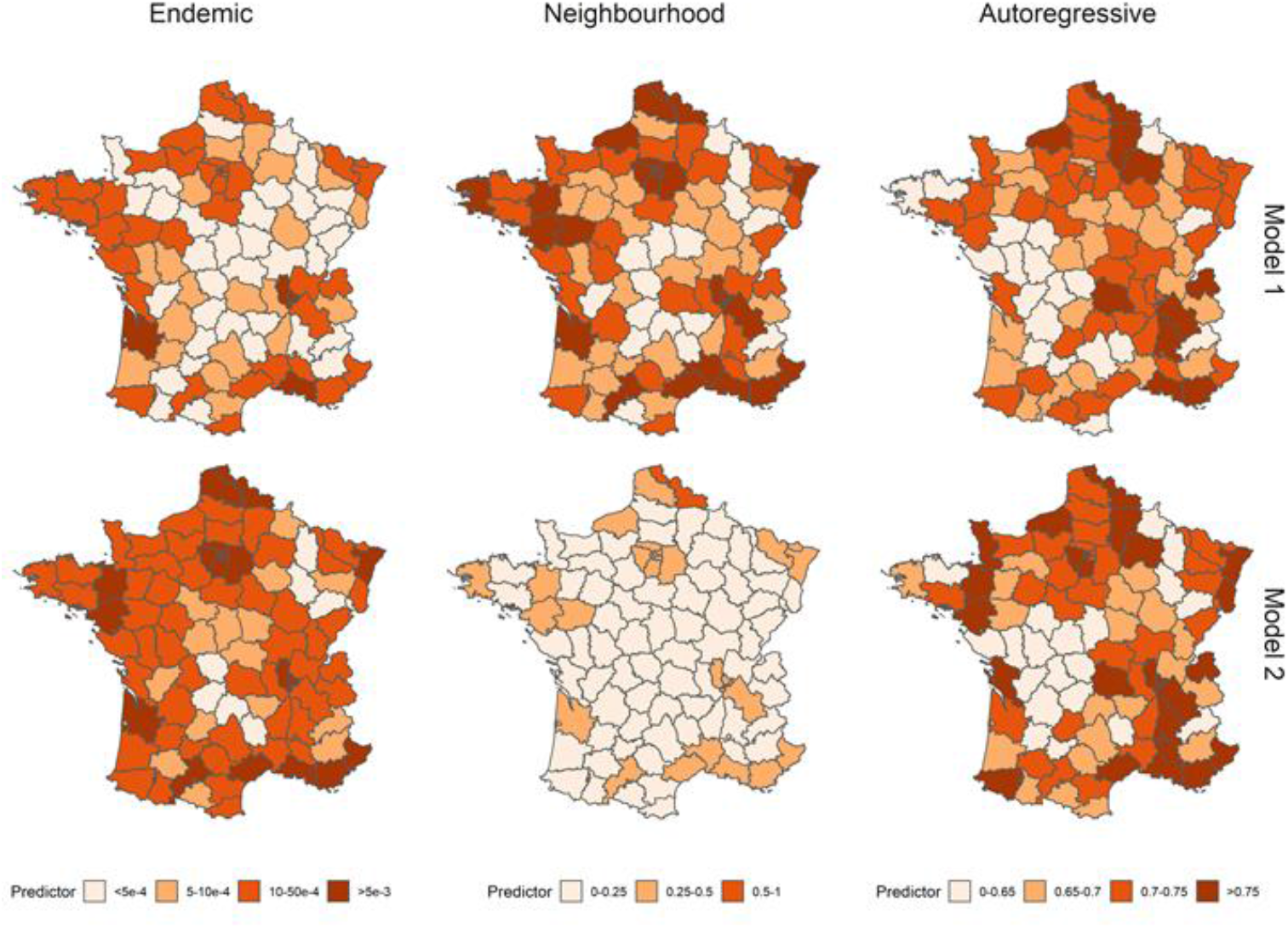
Average values of the endemic, neighbourhood, and autoregressive predictors per department in Model 1 (upper row) and Model 2 (lower row) over the year 2019. Since the absolute values are expected to vary over the year because of seasonality, the panels show the relative geographical heterogeneity. The endemic predictor corresponds to the number of importations per day per department, whereas the autoregressive predictor corresponds to the number of secondary cases per case in each department. The absolute value of the neighbourhood predictor is harder to interpret directly since it is multiplied by the connectivity matrix in the equation. Higher values were associated with departments with higher risks of observing cases following population movements.

As expected, the overall number of baseline importations in Model 1 was lower than in Model 2, which was compensated by a higher number of cross-regional transmissions (Figure 3). This shows that some of the cases that could not be linked to local transmission, or transmission between neighbouring departments in Model 2, were classified as cross-regional transmissions in Model 1, which would indicate long-distance transmission events. In both models, departments with a higher number of inhabitants were most at-risk of cross-regional and baseline importations, which corresponds to the strong association between the number of inhabitants and the endemic and neighbourhood components highlighted in Figure 2. Departments like Bouches-du-Rhône that combine a high number of inhabitants with low vaccine coverage were associated with the highest number of baseline and cross-regional importations in both models. The variations in the autoregressive component were smaller than in the importation-related components: For instance, the highest autoregressive predictor value (Var: 0.81 [0.74 - 0.88]) was 35% higher than the lowest value (Lozère: 0.60 [0.53 – 0.66]) in Model 1, whereas the number of baseline importations in Bouches-du-Rhônes was more than 100 times above the number of importations in Lozère (South of France). This can be explained by the coefficients of the autoregressive components being much closer to 0 than the most extreme coefficients in the importation-related components (Figure 2).

### Model fit and calibration

The daily and weekly fits of Model 1 and Model 2 indicate that they were able to match the transmission dynamics observed in France between 2009 and 2017, despite wide variations in the annual number of cases (Figure 4 Panel A and B, Supplement Section 4). In years where active transmission was reported, most of the cases stemmed from the autoregressive component, indicating that the local outbreaks were sustained by transmission within the departments. Indeed, across all years, the autoregressive component accounted for 72.9% of the cases, whereas 23.7% of the cases came from cross-regional transmission, and 3.4% from the endemic component (Supplement Figure S12). This shows that in Model 1, 97.6% of the cases were explained by the transmission stemming from other cases reported in the dataset (93.2% in Model 2). The endemic component described the minority of isolated cases that could not be linked to any concurrent transmission cluster. Therefore, these cases would be more likely to be reported at times of low national levels of transmission when no other case could be linked to them, which explains the shift in seasonality of the endemic component observed in Figure 2 and Supplement Section 3.

**Figure 4:**
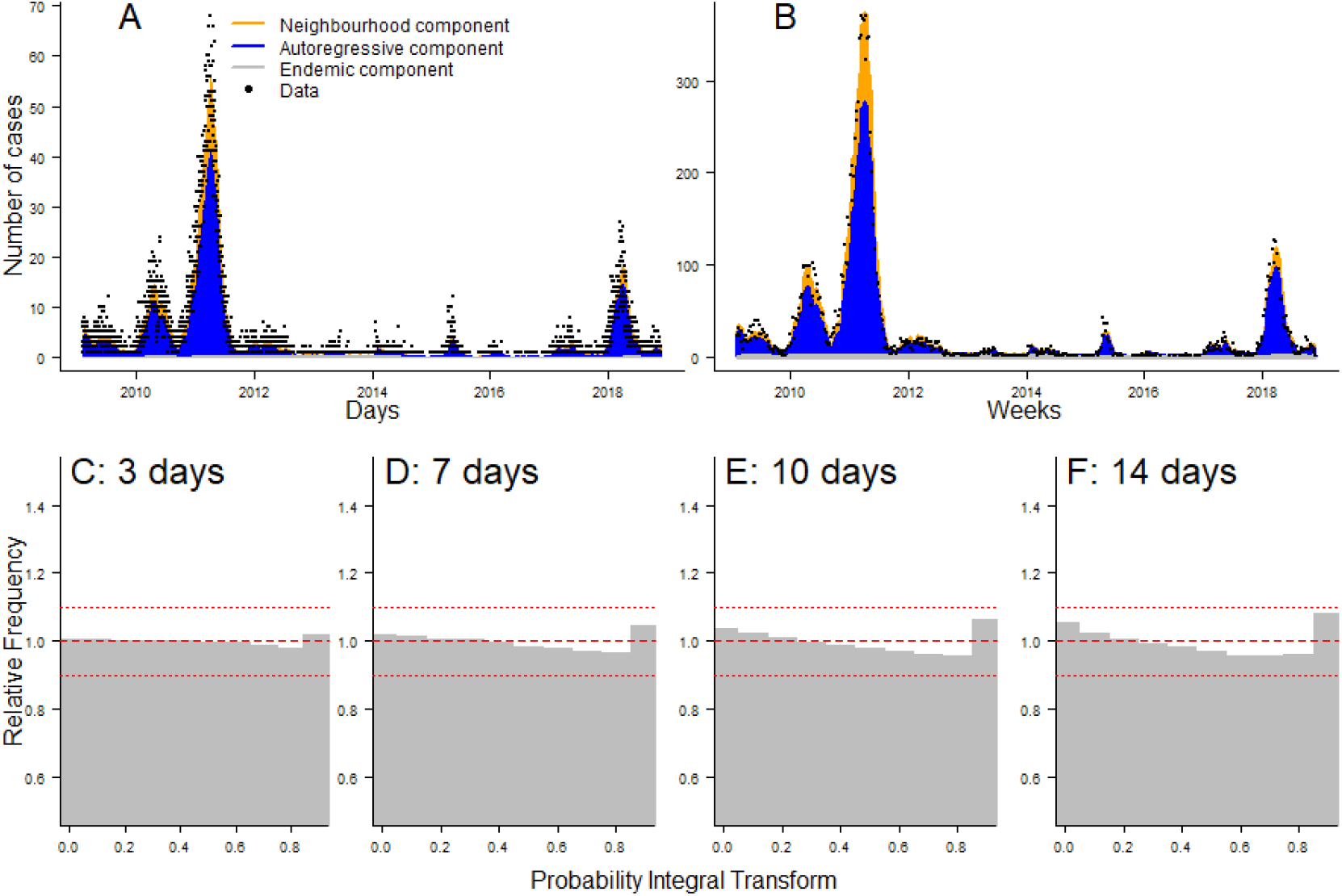
Panel A and B: Daily and weekly fit between the data and Model 1. The inferred number of cases is split among the three components of the model. Panel C to F: PIT histograms of Model 1, generated respectively for predictions 3, 7, 10, and 14 days ahead.

In order to visually assess the calibration of the model, and its ability to provide reliable short-term predictions for the number of cases per department, we generated PIT histograms showing the probability integral transform obtained when forecasting the number of cases 3, 7, 10, and 14 days ahead (Figure 4, Panels C to F). The PIT histogram is uniform for predictions 3 and 7 days ahead (all groups are above 0.9 and below 1.1), which shows the number of occurrences where the predictions of the model did not capture the number of cases one week ahead was not higher than expected under a uniform distribution. As we increased the number of days of forecast, there were more occurrences of the model mis-predicting the number of cases to come. Indeed, the U-shape observed in Panel F of Figure 4 indicates the model was less capable of identifying extreme events two weeks in advance. The calibration study indicated that Model 2 was more prone to under-estimating the number of cases than Model 1, and showed signs of bias for the 7, 10, and 14-day predictions (Supplement Section 4). The national number of cases predicted by Model 1 and Model 2 were similar, and match the data for predictions 7 days ahead (Supplement Figure S11). The AIC scores and the calibration study indicated Model 1 was able to fit the data better than Model 2 and was better calibrated. The rest of the Results section therefore focuses on the conclusions reached using Model 1. The equivalent analysis run on Model 2 is presented in the Supplementary Section 4.

### Impact of vaccination and recent incidence on onwards transmission

In order to illustrate the impact of recent outbreaks and variations in vaccine coverage on the transmission dynamics in France, we generated 10,000 simulations and computed the number of cases per department in 2019. We ran the simulation from August 2018 (during the historically low transmission season), until 31^st^ December 2019. We generated four sets of simulations under different initial conditions: using the last measures of average local vaccine coverage, category of recent incidence, and number of inhabitants; increasing or decreasing the vaccine coverage by three percent, and setting the category of recent incidence to 0 in each department.

Under the latest measures of coverage and incidence, the simulated outbreaks display a wide variation in the number of cases in 2019 (minimum 100 cases, median 1,100 cases, maximum 11,100 cases). Active transmission was generated in a wide range of departments. Indeed, across the simulation set, 44 of the 94 French departments reported more than 10 cases in at least 25% of the simulations. There was noteworthy spatial heterogeneity in the levels of incidence. Indeed, in 12 departments, there was no case generated in more than half of the simulations (Figure 5, top right panel). The departments most vulnerable to active transmissions were highly populated urban areas, such as Paris, the Bouches-du-Rhône, and the North of France. Because they are highly populated, these departments were susceptible to repeated importations (they reported at least 1 case in more than 95% of the simulations), which could then cause large transmission clusters. This was especially evident in the South-East of France, where we highlighted that the number of secondary cases per case in the department was among the highest in the country (Figure 3 and Figure 5). Numerous departments were affected by large outbreaks in a subset of the simulated datasets: 27 departments reported more than 50 cases in at least 5% of the simulations (Figure 5). Further, at least one major outbreak was generated in the majority of the simulations: in 55% of the simulations, one department reported more than 100 cases (the most commonly affected department were Paris and its surroundings, the Nord, and Bouches-du-Rhône).

**Figure 5:**
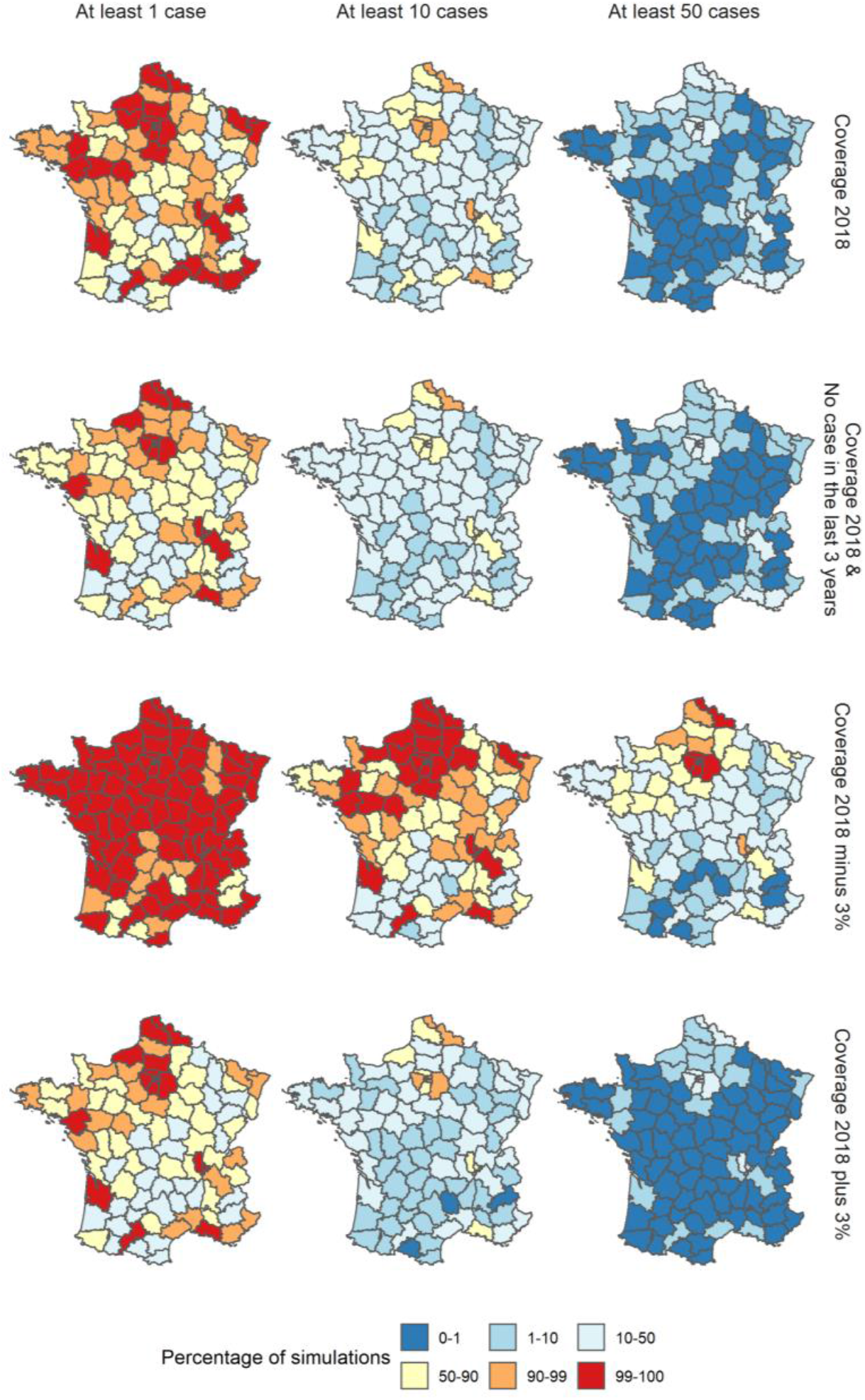
Percentage of simulations where the number of cases reported in each department in 2019 was at least 1, 10, and 50 cases for each scenario using parameter estimates from Model 1. Each row corresponds to a different scenario: i) Reference, ii) Minimum level of recent incidence in each department, iii) Local vaccine coverage decreased by three percent in each department, iv) Local vaccine coverage increased by three percent in each department.

Decreasing the average three-year vaccine coverage by three percent led to an important increase in the number of cases per outbreak (median 4,900 cases, more than 95% of the simulations resulted in more than 1,000 cases). This was first due to an increase in importations and cross-regional transmission: all 94 departments had at least one case in more than half of the simulations, 77 in at least 90% of the simulations. Furthermore, the decrease in vaccination coverage resulted in higher chances of uncontrolled transmissions in many departments (Figure 5, third row). On the other hand, increasing the vaccine coverage by three percent caused an important drop in the number of cases (median 605 cases, 80% of the simulations generated less than 1,000 cases), caused by both a decrease in the number of importations, and in the potential for secondary transmission following importations. Although outbreaks were still punctually generated, these events are much rarer than in the other two simulation sets: in 25.8% of the simulations, at least one department generated more than 100 cases (54.1% with the baseline scenario, 95.4% when we reduced the local vaccine coverage).

Finally, setting the local recent incidence to the minimum level in each department, which would fulfil the elimination guidelines, had two opposite effects: it led to a decrease in the number of importations and cross-regional transmission, and an increase in the number of infections within each department (Figure 2). In this simulation set, the number of departments where no cases were generated in more than half of the simulations was similar to when the vaccine coverage was increased (24 departments in this simulation set, 29 when the vaccine coverage was increased, Figure 5), which shows the reduction in the number of cross-regional transmission and background importations. Conversely, the number of large outbreaks was only marginally inferior to the reference simulation set: in 44% of the simulations, there were more than 100 cases generated in at least one department (54% in the reference dataset). The geographical distribution of the risks of large outbreaks was almost identical to the reference simulation set (Figure 5). Therefore, although the number of importations was reduced, changing the level of recent incidence did not have a clear impact on the risks of active transmission. More departments became vulnerable to secondary transmission, and despite importations in these departments being rarer, they were more likely to lead to large outbreaks when they happened. The two opposing effects recent incidence had on importation and transmission therefore created a different dynamic of transmission observed in the simulation set, without strongly reducing the risks of outbreaks.

Each of these simulation sets highlighted the wide range of scenarios that could be generated using the parameter distributions inferred by our model. In order to gain more understanding on the spatial spread and consequences of importations, we then explored the impact of localised repeated importations on overall transmission.

### Impact of local clusters of transmission

Since the endemic component, which can be interpreted as external importations, represented a minority of the cases in our model (Supplement Figure S12), repeated importations in a given department over a short timespan rarely occurred in the simulations. Furthermore, due to the seasonality of the endemic component, fewer importations are generated early in December to February, which corresponds to the peak period of the other components, and would therefore be more likely to cause secondary transmissions (Supplement Section 3). We simulated one year of transmission following ten importations in December 2018 to illustrate: i) the potential for local outbreaks, and ii) the spatial spread of transmission following repeated local importations. We selected four departments to compare the impact of repeated importations in a range of settings: Paris (many inhabitants, 91% vaccine coverage, surrounded by urban areas), Bouches-du-Rhône (many inhabitants, 84% vaccine coverage), Haute Garonne (many inhabitants, 91% vaccine coverage but high levels of recent incidence, surrounded by rural areas with lower vaccine coverage), and Gers (Rural area, 79% vaccine coverage) (Figure 6).

**Figure 6:**
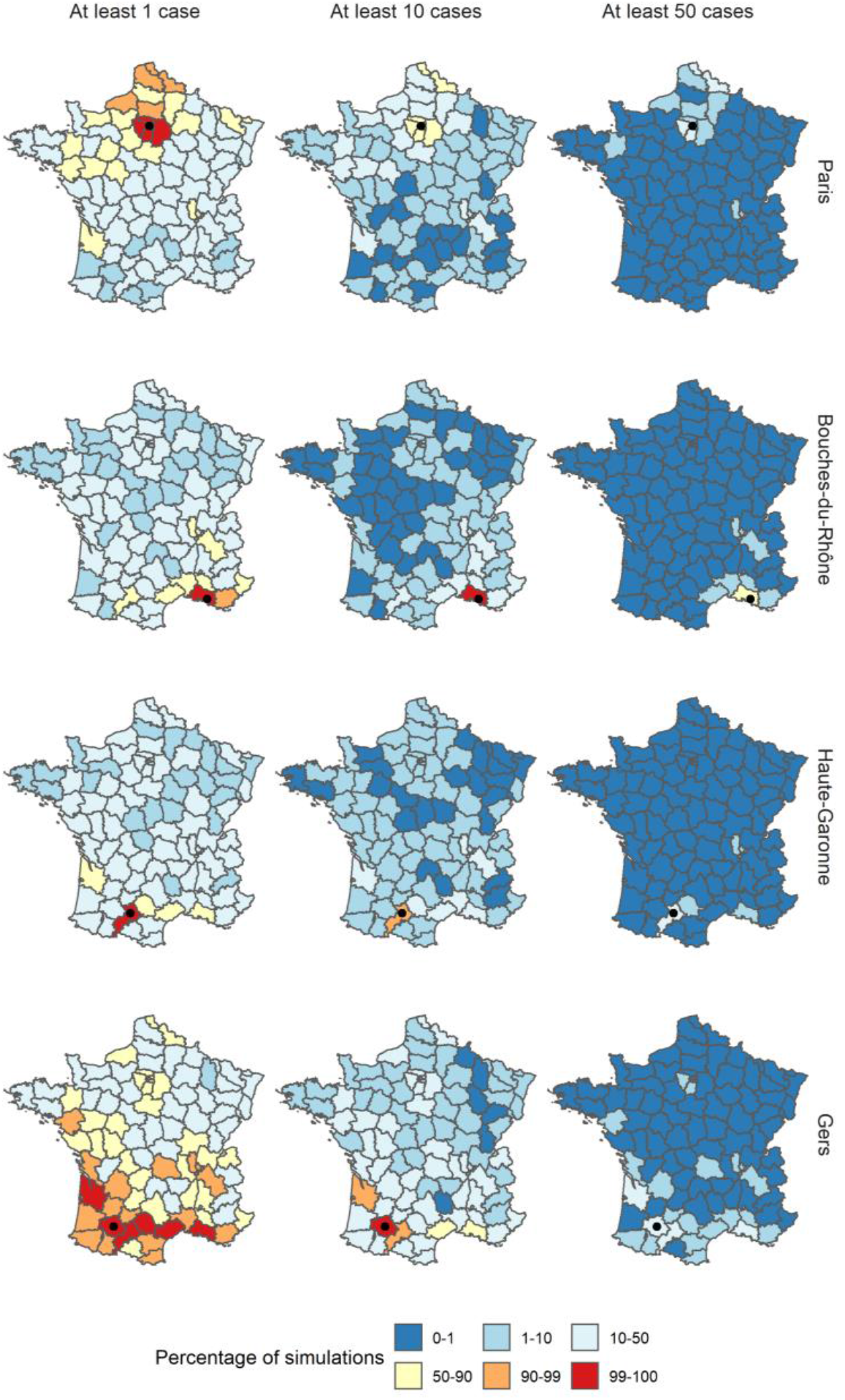
Percentage of simulations where the number of cases reported in each department in 2019 was at least 1, 10, and 50 cases following the importations of ten cases in December 2018, and using the parameter estimates from Model 1. For each row, the department of importation is indicated by a black dot.

Firstly, major local outbreaks in the department of importation were generated in all four simulation sets, and especially in Paris and Bouches-du-Rhône, where the proportion of simulations that yielded more than 100 subsequent cases in the department was 40% and 39%, respectively. In the Bouches-du-Rhône, large outbreaks were mostly due to the low vaccination coverage, whereas in Paris, outbreaks were mostly linked to the connectivity to nearby areas and the high number of inhabitants, which meant the department was likely to attract cross-regional transmissions. Major local outbreaks were rarer in the other two scenarios (9% of simulations above 100 in Haute Garonne, 10% in Gers). The lower proportion of large outbreaks resulted from different factors: recent large outbreaks in Haute Garonne reduced the autoregressive predictor, lowering the number of secondary cases per case imported; whereas since Gers is a rural department, with a low number of inhabitants, almost all the local cases were due to local transmission (auto-regressive component), with very few cross-regional transmissions into Gers.

Conversely, the simulations where cases were imported in Gers yielded the largest spatial spread throughout the country: the median number of departments that reported at least 1 case was 53 (16 when the importations were generated in Haute Garonne; 15 in Bouches-du-Rhône; 39 in Paris). As stated in the method, the number of cross-regional transmissions is the product of the predictor and the connectivity matrix, divided by the number of inhabitants in the department of origin, to represent that only a fraction of commuters will be infected. Therefore, populous areas are more likely to attract cross-regional transmissions, whereas more rural departments are more likely to seed outbreaks in other areas. The relatively high spatial spread when cases were imported in Paris is due to the short distance between Paris and its suburbs, which is then more likely to cause cross-regional transmission in the northern departments. Despite the cross-regional spread observed in both of these simulations sets, outbreaks remained local, and occurrences of nation-wide outbreaks were almost null. The departments most at risk of outbreak following cross-regional spread were some of the direct neighbours of the department of importations, or the large urban areas (Figure 6). To further explore this, we ran the same simulations decreasing the vaccine coverage by three percent, which greatly increased the number of departments exposed in each simulation set, and increased the risk of local transmission (Supplement Section 6). Therefore, although repeated importations could cause active transmission in and around the departments of importation, the current values of vaccine coverage and the seasonality of transmission were able to prevent nationwide transmission.

## Discussion

This analysis explored which local factors were associated with high risks of transmission in France over the last decade. Since 2017, immunity gaps, caused by failures to vaccinate, have been linked to a resurgence of measles in all WHO regions [38]. In countries near-elimination, large outbreaks have been linked to heterogeneity in the levels of immunity, with pockets of susceptibles fuelling punctual outbreaks despite high national vaccine uptake [1,2,4,25]. Our study showed that local values of vaccine coverage were linked to lower transmission, whereas lower levels of recent incidence were not associated with lower risks of local transmission. Furthermore, we highlighted that a drop of 3% in the three-year vaccine coverage triggered a five-fold increase in the number of cases simulated in a year.

The fact that higher vaccine coverage was associated with a lower number of secondary cases is consistent with prior expectations, and would confirm that the local values of first dose vaccine coverage are a good indicator of the actual immunity in the population and risks of future transmission. Reporting accurate values of local vaccine coverage is challenging, for instance because the vaccination status of people moving regions can be hard to track and lead to measurement errors. Furthermore, we did not have access to complete data on the coverage of the second MMR dose, which would be a better indicator of vulnerable areas. Therefore, detecting the association between recent vaccine uptake and incidence is encouraging. The impact of local vaccination coverage on transmission may also be muddled by sub-regional vaccine heterogeneity. For instance, pockets of susceptibles within a region, i.e. areas within the region where the vaccine coverage is substantially lower than the regional average, may be at high risk of transmission and would not be observable in regional coverage [39]. This phenomenon can only be hypothesised here, and could be explored using local data on incidence and vaccine uptake at a sub-regional scale.

Variations in vaccine coverage had a noticeable impact on the number of cases generated in the simulation study. We showed the effects of a three percent increase and decrease of the three-year average vaccine coverage on the number of cases, which highlighted the risks of uncontrolled transmission in the event of a decrease of vaccine-induced protection. Events such as the disruption caused by the SARS-COV-19 pandemic on routine measles vaccination campaigns could therefore highly increase the risks of uncontrolled measles transmission in the years to come [40,41].

The departments that reported few cases per million in the past three years were associated with higher risks of local transmission (autoregressive component). Therefore, according to our model, regions eligible for elimination status were not associated with lower risks of onwards transmission. Conversely, high levels of recent transmission were associated with a lower number of cross-regional transmissions and importations, although we cannot methodologically establish the causality of this association. The impact on the simulation study was clear: when we set the category of recent incidence to the lowest level, departments were less exposed to cases, and spatial spread was rarer, whilst there was little change in the risks of major outbreaks. The simulations showed an ‘all-or-nothing’ situation: departments tended to report very few to no cases, whilst also being more likely to be affected by outbreaks. These results would indicate that looking into the level of incidence to quantify the future risks of outbreaks can be deceptive, and importations in a department with low recent incidence would result in large transmission clusters.

We proposed a new framing of the Epidemic-Endemic model implemented in *hhh4* by adapting it to daily count data using the distribution of the serial interval to compute the local transmission potential. Using daily case counts allowed us to avoid biases associated with aggregated case counts, such as the influence of the arbitrary aggregation date, by accounting for the impact of variation in the serial intervals. We also accounted for the risks of unreported cases by computing a composite multimodal serial interval, thus allowing for transmission with a missing generation, or an unreported ancestor. The model was able to capture the dynamic of transmission better than the 10-day aggregated model, as shown by the calibration study (Supplement Section 7). Nevertheless, our framing of the *hhh4* model introduced new biases: we used a distribution of the serial interval based on previous studies rather than estimating the weights during the fitting procedure and set the proportion of missing generations in the composite serial interval. We explored the impact of the proportion of missing generations by fitting the model with different composite serial intervals and concluded that the impact of each covariate was robust to these changes (Supplement Section 1). We also integrated a potential day-of-the-week effect, and observed that although it had an impact on the auto-regressive component, it did not change the estimates of the other parameters, and therefore did not change the conclusions of the study (Supplement Section 8).

Using the *hhh4* model allowed us to analyse the different impact of various covariates on local and cross-regional transmission, and background importation of cases. According to the models we implemented, an overwhelming majority (>90%) of the transmission came from the cross-regional and local components of the regression. This indicates that in the models, the endemic component only corresponds to rare background cases that could not be linked to concurrent transmission events. This could point towards model misspecifications, for example, connecting unrelated importations to concurrent local transmission. Since endemic transmission tends to refer to cases otherwise unexplained by the mechanistic components, the seasonality of the endemic component is decoupled from the other components, i.e. endemic cases are likely when local and cross-regional transmission are lower.

Since the endemic component accounted for such a small minority of the cases, group importations of cases in a given department were rarely observed in the simulations. However, tourism, and local events lead to large gatherings and can increase the risks of group importations in a limited period of time [36,37]. We simulated the spatial spread following repeated importations in a given department, and highlighted that although large outbreaks in the department of importations were common, nation-wide transmission following these importations was very rare. Only the departments where all cases had been imported, and its neighbours, were at risk of uncontrolled outbreaks. Decreasing the level of vaccination by three percent was associated with a large increase in the level of exposure of all departments, and in the number of departments where large outbreaks were generated (Supplement Section 6 and 7). The high levels of transmission observed in recent years in France suggest that importations are frequent, and even a small drop in vaccination could dramatically increase measles transmission in the country.

Furthermore, since the number of inhabitants was strongly associated with risks of background importations, most of the endemic importations were reported in urban areas, where the risks of exportations were lower. This could explain the discrepancies between the distribution of the number of cases in the simulations (Figure 5, top row), and the actual number of cases reported in France in 2019 [42]. Active transmission was reported in a number of rural areas, notably in the South West of France, and in Savoie (East). This could be due to importations and cross-regional transmission that are under-estimated by our model. Although the model captured the dynamics seen in the data, the calibration study showed it was only able to predict short-term transmission up to one week. The PIT histogram associated to the 14-day calibration displayed signs of bias, which shows that the model was not able to consistently predict variations in the future number of cases in the next two weeks. We identify several factors that could explain the discrepancies observed for longer term predictions: i) the indicator of local immunity we used was flawed: two-dose coverage would be a better indicator of the proportion of the population that is protected; ii) The sub-regional heterogeneity in coverage and past incidence within the department that could be concealed by NUTS3 aggregated data: because of social groups that rarely mix with one another, or large NUTS regions, large outbreaks in a given community would not be a good indicator of the overall level of immunity in a region. Nevertheless, we believe that the results obtained using limited publicly available covariates are encouraging and we intend to apply this method using more complete data.

We identified a number of limitations of this study that have not yet been mentioned: Firstly, potential reactive control measures in case of high transmission were not accounted for. It is likely that if the level of incidence was increasing over a short period of time, control measures would be implemented and the behaviour of the individuals may change (e.g. school closures, catch-up vaccination campaigns). This could impact the number of expected cases after a certain threshold is passed, and impact the dynamics in the simulated outbreaks. Secondly, we did not include information on the age or genotype of the cases. Therefore, unrelated importations in successive time-steps in a given region may be considered as linked by our model, whereas they should be separated. Further development of this method could focus on taking this aspect into account, in order to give information on the number of independent concurrent chains. Thirdly, since this is not a transmission model, some extreme values could trigger unlikely behaviour. For instance, if the vaccination rate would be 100%, we would still expect sporadic transmission. Although this would not be entirely implausible given that only the vaccination coverage in the past three years was taken into account in the models (i.e. even if it was 100% coverage, there could be susceptible individuals in different age groups). Finally, the impact of the different covariates on the number of cases was constant through time. For instance, the impact of seasonality may depend on factors such as the weather which may vary each year, which would not be accounted for in the model we developed.

We used variables collected in a wide range of settings (regional vaccine coverage, incidence, number of inhabitants, surface), therefore this analysis can be reproduced in other countries to analyse the potential for local transmission as well as the impact of recent incidence and vaccine-induced immunity. Since the case counts data are not publicly available, we share the code used to generate the analysis applied to a simulated dataset on a Github repository: (https://github.com/alxsrobert/measles-regional-transmission).

## Data Availability

The daily case counts data came from the European Surveillance System (TESSy), provided by France and released by ECDC. The data cannot be shared publicly. To make this study as reproducible as possible, we generated simulated case counts in France over the same timespan as the main analysis. The code used to generate the simulated dataset, and all the figures presented in the paper is shared in the Github repository https://github.com/alxsrobert/measles-regional-immunity, which is attached to this paper. This repository also contains the publicly available covariates used in the model.

## Data availability

The daily case counts data came from the European Surveillance System – TESSy, provided by Santé Publique France and released by ECDC. The data cannot be shared publicly. To make this study as reproducible as possible, we generated simulated case counts in France over the same timespan as the main analysis. The code used to generate the simulated dataset, and all the figures presented in the paper is shared in a Github repository (https://github.com/alxsrobert/measles-regional-transmission). This repository also contains the publicly available covariates used in the model (local vaccine coverage, number of inhabitants, surface, distance between departments).

## Disclaimer

The views and opinions of the authors expressed herein do not necessarily state or reflect those of ECDC. The accuracy of the authors’ statistical analysis and the findings they report are not the responsibility of ECDC. ECDC is not responsible for conclusions or opinions drawn from the data provided. ECDC is not responsible for the correctness of the data and for data management, data merging and data collation after provision of the data. ECDC shall not be held liable for improper or incorrect use of the data.

## Acknowledgments

We acknowledge Helen Johnson and Nick Bundle from ECDC, who participated in the analysis plan. We also acknowledge the ECDC and Santé Publique France for collecting and providing the case count data used in this study.

## Author Contributions

AR, SF and AJK developed the method and the analysis plan. AR implemented the analysis, wrote the code and ran the model. AR interpreted the results, with contributions from SF and AJK. AR wrote the first draft and the supplementary material. AR, SF, AJK contributed to the manuscript, all authors approved the final version.

## References

[1] Gastañaduy PA, Budd J, Fisher N, Redd SB, Fletcher J, Miller J, et al. A Measles Outbreak in an Underimmunized Amish Community in Ohio. N Engl J Med 2016;375:1343–54. doi:10.1056/NEJMoa1602295.

[2] Woudenberg T, Van Binnendijk RS, Sanders EAM, Wallinga J, De Melker HE, Ruijs WLM, et al. Large measles epidemic in the Netherlands, May 2013 to March 2014: Changing epidemiology. Eurosurveillance 2017;22:1–9. doi:10.2807/1560-7917.ES.2017.22.3.30443.

[3] Funk S, Knapp JK, Lebo E, Reef SE, Dabbagh AJ, Kretsinger K, et al. Combining serological and contact data to derive target immunity levels for achieving and maintaining measles elimination. BMC Med 2019. doi:10.1186/s12916-019-1413-7.

[4] Glasser JW, Feng Z, Omer SB, Smith PJ, Rodewald LE. The effect of heterogeneity in uptake of the measles, mumps, and rubella vaccine on the potential for outbreaks of measles: A modelling study. Lancet Infect Dis 2016;16:599–605. doi:10.1016/S1473-3099(16)00004-9.

[5] Keenan A, Ghebrehewet S, Vivancos R, Seddon D, MacPherson P, Hungerford D. Measles outbreaks in the UK, is it when and where, rather than if? A database cohort study of childhood population susceptibility in Liverpool, UK. BMJ Open 2017;7. doi:10.1136/bmjopen-2016-014106.

[6] World Health Organization (WHO). Global Vaccine Action Plan Global Vaccine Action Plan. Who 2011:4–7.

[7] World Health Organization. Framework for verifying elimination of measles and rubella. Wkly Epidemiol Rec 2013;88:89–100. doi:10.1371/jour.

[8] World Health Organization (WHO). European Region loses ground in effort to eliminate measles 2019.

[9] Pan American Health Organization. Epidemiological Update: Measles. Paho/ Who 2019;2020:1–12.

[10] Fraser B. Measles outbreak in the Americas. Lancet (London, England) 2018;392:373. doi:10.1016/S0140-6736(18)31727-6.

[11] Litvoc MN, Lopes MIBF. From the measles-free status to the current outbreak in Brasil. Rev Assoc Med Bras 2019;65:1229–30. doi:10.1590/1806-9282.65.10.1129.

[12] Dimala CA, Kadia BM, Nji MAM, Bechem NN. Factors associated with measles resurgence in the United States in the post-elimination era. Sci Rep 2021;11:1–10. doi:10.1038/s41598-020-80214-3.

[13] Bernadou A, Astrugue C, Méchain M, Le Galliard V, Verdun-Esquer C, Dupuy F, et al. Measles outbreak linked to insufficient vaccination coverage in Nouvelle-Aquitaine region, France, October 2017 to July 2018. Eurosurveillance 2018;23:1–5. doi:10.2807/1560-7917.ES.2018.23.30.1800373.

[14] Held L, Höhle M, Hofmann M. A statistical framework for the analysis of multivariate infectious disease surveillance counts. Stat Modelling 2005;5:187–99. doi:10.1191/1471082X05st098oa.

[15] Meyer S, Held L, Höhle M. hhh4: Endemic-epidemic modeling of areal count time series. J Stat Softw 2016.

[16] Lloyd-Smith JO, Schreiber SJ, Kopp PE, Getz WM. Superspreading and the effect of individual variation on disease emergence. Nature 2005;438:355–9. doi:10.1038/nature04153.

[17] Fine PEM. The Interval between Successive Cases of an Infectious Disease. Am J Epidemiol 2003;158:1039–47. doi:10.1093/aje/kwg251.

[18] Bjørnstad ON, Finkenstädt BF, Grenfell BT. Dynamics of measles epidemics: Estimating scaling of transmission rates using a Time series SIR model. Ecol Monogr 2002;72:169–84. doi:10.1890/0012-9615(2002)072[0169:DOMEES]2.0.CO;2.

[19] Bracher J, Held L. Endemic-epidemic models with discrete-time serial interval distributions for infectious disease prediction. Int J Forecast 2020. doi:10.1016/j.ijforecast.2020.07.002.

[20] Woudenberg T, Woonink F, Kerkhof J, Cox K, Ruijs WLM. The tip of the iceberg : incompleteness of measles reporting during a large outbreak in The Netherlands in 2013 – 2014. Epidemiol Infect 2018;146:716–22. doi:https://doi.org/10.1017/S0950268818002698.

[21] Lenormand M, Bassolas A, Ramasco JJ. Systematic comparison of trip distribution laws and models. J Transp Geogr 2016;51:158–69. doi:10.1016/j.jtrangeo.2015.12.008.

[22] Institut National de la Statistique et des Etudes Economiques. Estimation de la population au 1er janvier 2020 2020. https://www.insee.fr/fr/statistiques/1893198#consulter (accessed September 7, 2020).

[23] Eurostat. European population grid cells 2011. https://ec.europa.eu/eurostat/web/gisco/geodata/reference-data/grids (accessed September 12, 2020).

[24] Hijmans RJ, Etten J van, Mattiuzzi M, Sumner M, Greenberg JA, Lamigueiro OP, et al. Package “raster.” R 2014.

[25] Herzog SA, Paul M, Held L. Heterogeneity in vaccination coverage explains the size and occurrence of measles epidemics in German surveillance data. Epidemiol Infect 2011;139:505–15. doi:10.1017/S0950268810001664.

[26] Santé Publique France. Données départementales 2007-2012 de couverture vaccinale rougeole, rubéole, oreillons à 24 mois 2019. https://www.santepubliquefrance.fr/determinants-de-sante/vaccination/articles/donnees-departementales-2007-2012-de-couverture-vaccinale-rougeole-rubeole-oreillons-a-24-mois (accessed September 7, 2020).

[27] Santé Publique France. Estimations des couvertures vaccinales à 24 mois à partir des certificats de santé du 24e mois, 2004-2007 2010. https://www.santepubliquefrance.fr/determinants-de-sante/vaccination/articles/donnees-departementales-2013-2017-de-couverture-vaccinale-rougeole-rubeole-oreillons-a-24-mois (accessed September 7, 2020).

[28] Santé Publique France. Données départementales 2013-2017 de couverture vaccinale rougeole, rubéole, oreillons à 24 mois 2019. https://www.santepubliquefrance.fr/determinants-de-sante/vaccination/articles/donnees-departementales-2013-2017-de-couverture-vaccinale-rougeole-rubeole-oreillons-a-24-mois (accessed September 7, 2020).

[29] Antona D, Lévy-Bruhl D, Baudon C, Freymuth F, Lamy M, Maine C, et al. Measles elimination efforts and 2008-2011 outbreak, France. Emerg Infect Dis 2013;19:357–64. doi:10.3201/eid1903.121360.

[30] Institut de Veille Sanitaire. Données de déclaration obligatoire de la rougeole. 2009.

[31] Fitzpatrick G, Ward M, Ennis O, Johnson H, Cotter S, Carr MJ, et al. Use of a geographic information system to map cases of measles in real-time during an outbreak in Dublin, Ireland, 2011. Eurosurveillance 2012;17:1–11. doi:10.2807/ese.17.49.20330-en.

[32] Yang W, Wen L, Li SL, Chen K, Zhang WY, Shaman J. Geospatial characteristics of measles transmission in China during 2005−2014. PLoS Comput Biol 2017;13:1–21. doi:10.1371/journal.pcbi.1005474.

[33] Andrianou XD, Del Manso M, Bella A, Vescio MF, Baggieri M, Rota MC, et al. Spatiotemporal distribution and determinants of measles incidence during a large outbreak, Italy, september 2016 to july 2018. Eurosurveillance 2019;24:1–12. doi:10.2807/1560-7917.ES.2019.24.17.1800679.

[34] Funk S, Camacho A, Kucharski AJ, Lowe R, Eggo RM, Edmunds WJ. Assessing the performance of real-time epidemic forecasts: A case study of Ebola in the Western Area Region of Sierra Leone, 2014–15. BioRxiv 2017:1–17. doi:10.1101/177451.

[35] Czado C, Gneiting T, Held L. Predictive Model Assessment for Count Data 2009:1254–61. doi:10.1111/j.1541-0420.2009.01191.x.

[36] le Polain de Waroux O, Saliba V, Cottrell S, Young N, Perry M, Bukasa A, et al. Summer music and arts festivals as hot spots for measles transmission: Experience from England and Wales, June to October 2016. Eurosurveillance 2016;21:1–6. doi:10.2807/1560-7917.ES.2016.21.44.30390.

[37] Gautret P, Steffen R. Communicable diseases as health risks at mass gatherings other than Hajj: What is the evidence? Int J Infect Dis 2016;47:46–52. doi:10.1016/j.ijid.2016.03.007.

[38] Patel MK, Goodson JL, Alexander JP, Kretsinger K, Sodha S V, Steulet C. Progress Toward Regional Measles Elimination — Worldwide, 2000 – 2019 2020;69:1700–5.

[39] Blumberg S, Enanoria WTA, Lloyd-Smith JO, Lietman TM, Porco TC. Identifying postelimination trends for the introduction and transmissibility of measles in the United States. Am J Epidemiol 2014;179:1375–82. doi:10.1093/aje/kwu068.

[40] Saxena S, Skirrow H, Bedford H. Routine vaccination during covid-19 pandemic response. BMJ 2020;369. doi:10.1136/bmj.m268.

[41] Dinleyici EC, Borrow R, Safadi MAP, van Damme P, Munoz FM. Vaccines and routine immunization strategies during the COVID-19 pandemic. Hum Vaccines Immunother 2021;17:400–7. doi:10.1080/21645515.2020.1804776.

[42] Santé Publique France. Bulletin épidémiologique rougeole. Données de surveillance 2019. 2020. https://www.santepubliquefrance.fr/content/download/231366/2508985 (accessed May 3, 2021).

