## Supplementary Material for "The impact of local vaccine coverage and recent incidence on measles transmission in France between 2009 and 2018"


### 1. Sensitivity analysis: Composite serial interval

In the main analysis, we considered that 50% of the composite serial interval reflected direct transmission (without missing generations between cases), and 50% came from the two scenarios with unreported cases. In order to analyse the impact of the proportion of direct transmission in the composite serial interval, we fitted Model 1 and Model 2 using different composite serial intervals, and reported the fitted distributions of the parameters. We computed ten different composite serial intervals with the proportion of direct transmission increasing from 10% to 100% by increments of 10%.

The impact of the covariates on the risks of transmission was robust to changes in the composite intervals for both Model 1 and Model 2 (Figure S1 and Figure S2). The median estimate of each parameter was included in the 95% confidence interval of the reference results (when the proportion of direct transmission in the composite serial interval is 50%). The only parameter that was impacted was the overdispersion parameter, which increased for most of the fits. This could indicate a wider difference between the mean estimate and the data, which would be taken into account by increasing the dispersion of the negative binomial distribution.

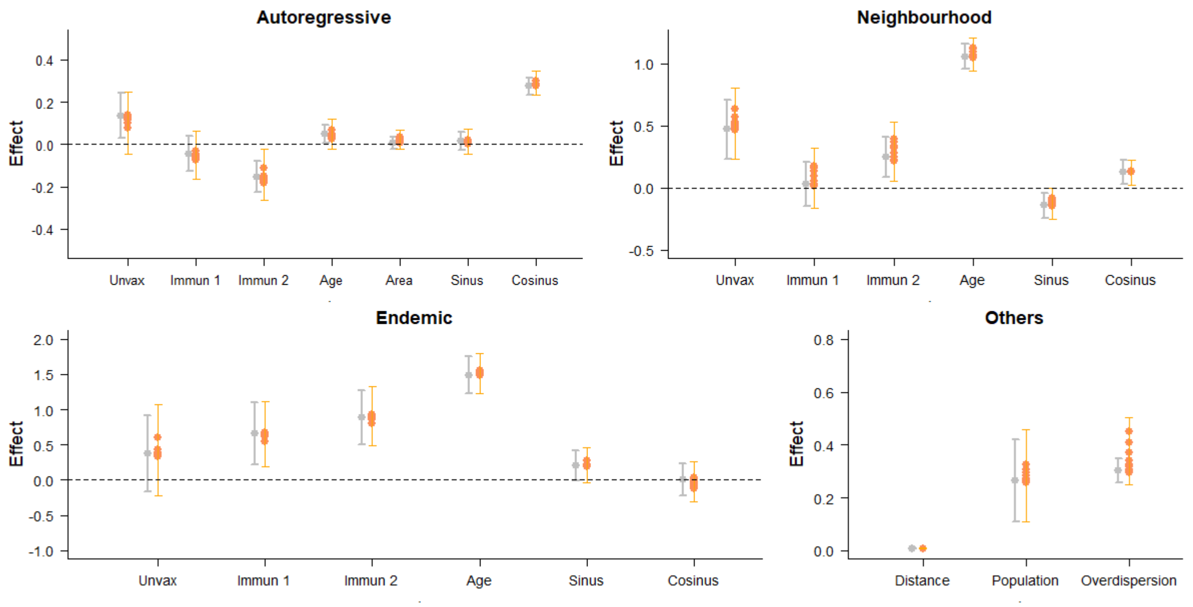


*Figure S1: Estimates of the parameters in each component of Model 1, using the reference fit (grey) and ten different values of the composite interval (orange). unvax corresponds to the effect of $u_{i,t}$, the mean proportion unvaccinated over the three years before t in i; incid1 and incid2 correspond to the effect of $N_{i,t}^{1}$ and $N_{i,t}^{2}$ the category of incidence in the three years before t in i; pop corresponds to the effect of $m_{i,t}$ the number of inhabitants at t in i; area corresponds to the effect of the surface; sin and cos correspond to the effects of seasonality; distance and population correspond to the spatial parameters of the connectivity matrix w (δ and γ); overdisp is the estimate of the log-overdispersion parameter in the negative binomial distribution of $Y_{i,t}$. Dots show the mean values associated with the parameters; arrows show the 95% Confidence interval. The orange arrows indicate the extreme values of the 95% confidence interval obtained using different distributions of the composite serial intervals.*

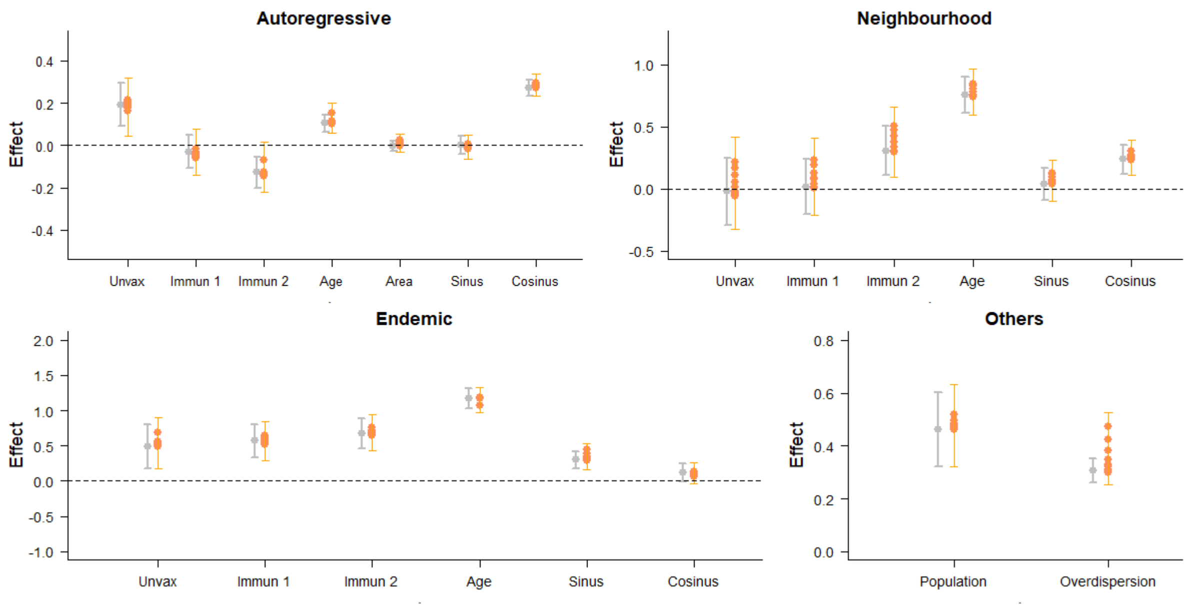


*Figure S2: Estimates of the parameters in each component of Model 2, using the reference fit (grey) and ten different values of the composite interval (orange). unvax corresponds to the effect of $u_{i,t}$, the mean proportion unvaccinated over the three years before t in i; incid1 and incid2 correspond to the effect of $N_{i,t}^{1}$ and $N_{i,t}^{2}$ the category of incidence in the three years before t in i; pop corresponds to the effect of $m_{i,t}$ the number of inhabitants at t in i; area corresponds to the effect of the surface; sin and cos correspond to the effects of seasonality; distance and population correspond to the spatial parameters of the connectivity matrix w (δ and γ); overdisp is the estimate of the log-overdispersion parameter in the negative binomial distribution of $Y_{i,t}$. Dots show the mean values associated with the parameters; arrows show the 95% Confidence interval. The orange arrows indicate the extreme values of the 95% confidence interval obtained using different distributions of the composite serial intervals.*

### 2. Inference of missing data in the regional vaccine coverage

We used publicly available data on 1st dose vaccine uptake between 2004 and 2017 in each department of metropolitan France to calculate the average local vaccine coverage over the past three years. There was no reported value of coverage reported in 2009, therefore the average in 2010, 2011 and 2012 were calculated using only two previous years. Besides 2009, there were 208 missing entries between 2006 and 2017 (17% of all entries), some regions had three years of consecutive unreported coverage, which made it impossible to compute an average without inferring the missing values. Since the incidence dataset starts in 2009, we did not infer the missing coverage data prior to 2006 (Figure S3).

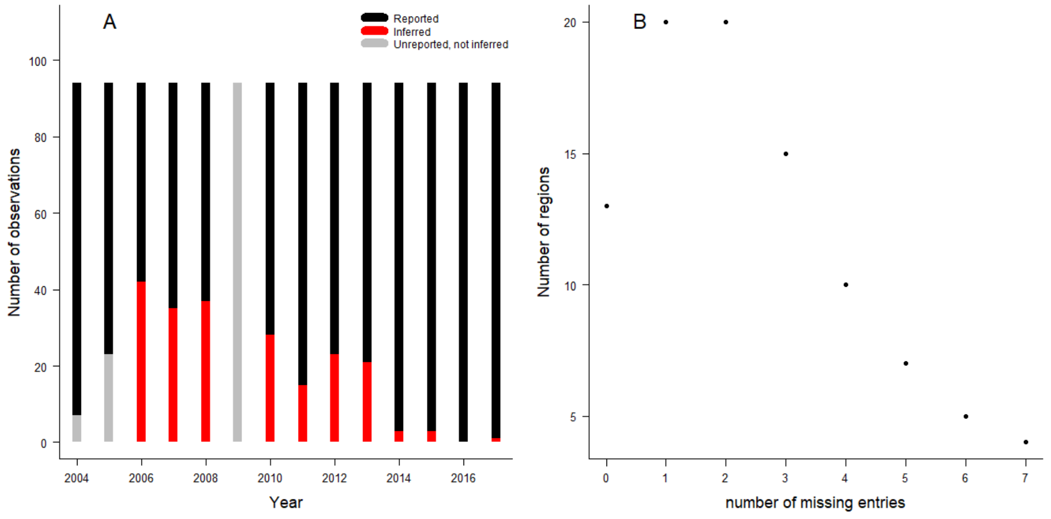


*Figure S3: Temporal and spatial distribution of the missing data: Panel A: Number of missing observations per year, missing coverage in 2004 and 2005 did not need to be inferred in the model, and since 2009 was entirely missing, we did not infer any value that year. Panel B: Number of missing entries per region.*

We implemented a beta mixed model to fit the annual local values of coverage. We used a beta regression since it is most adapted to modelling proportions or percentages. This model was implemented using the R package *glmmTMB* [1]. Observations are clustered over time within a region. The explanatory variables were orthogonal polynomials of degree 2 over the years covered by the data ($t$ varies between $1$ and $14$).

$$\text{logit}\,(Y_{ij}) = \beta_{0j} + \beta_{1j}t_i + \beta_{2j}t_i^2$$

Where:

$$\beta_{0j} = \gamma_{00} + U_{0j}$$
$$\beta_{1j} = \gamma_{10} + U_{1j}$$
$$\beta_{2j} = \gamma_{20} + U_{2j}$$

Using two-degree orthogonal polynomials gave more flexibility to the fitted curve than only using linear values of time. The regional values of intercept, and the impact of the orthogonal polynomial varied depending on the area, and were distributed around the fixed effect (Top panel of Figure S4). We show the average fitted trajectory of the vaccine coverage through time, along with the fit in three regions. This highlights that, although the average trajectory slightly increased between 2004 and 2017, there was no abrupt change in the first dose vaccine uptake. Nevertheless, using random effects allows for flexibility in other regions, whereby the fitted trajectories can show greater changes (e.g. region 2 in Figure S4).

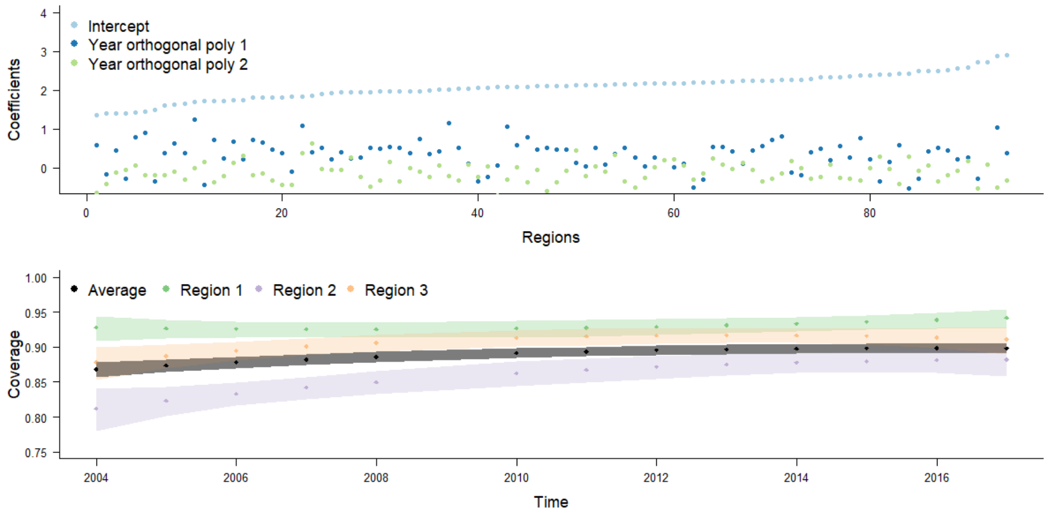


*Figure S4: Panel A: Values of the three parameters of the regression for each region. Panel B: Estimated values of coverage between 2004 and 2017, the dots represent the mean estimate, shaded areas correspond to the 95% Confidence Intervals. The purple, orange and green areas representing three departments illustrate how the changes through time can differ depending on the region.*

There was no discontinuous jump in the distribution of the random effect distribution for the three parameters of the model (Top panel of Figure S4). The fitted residuals plot did not show any clear trend relative to the dispersion of the data (Figure S5). We used different diagnostic tools provided by the R package *DHARMA* to test whether the model was correctly specified using simulated residuals. The outlier and the dispersion tests did not show any discrepancy, but the Kolgomorov Smirnov test of uniformity showed significant deviation from the expected distribution of residuals. This was expected from the minor discrepancies shown on the QQ plot (right panel of Figure S5).

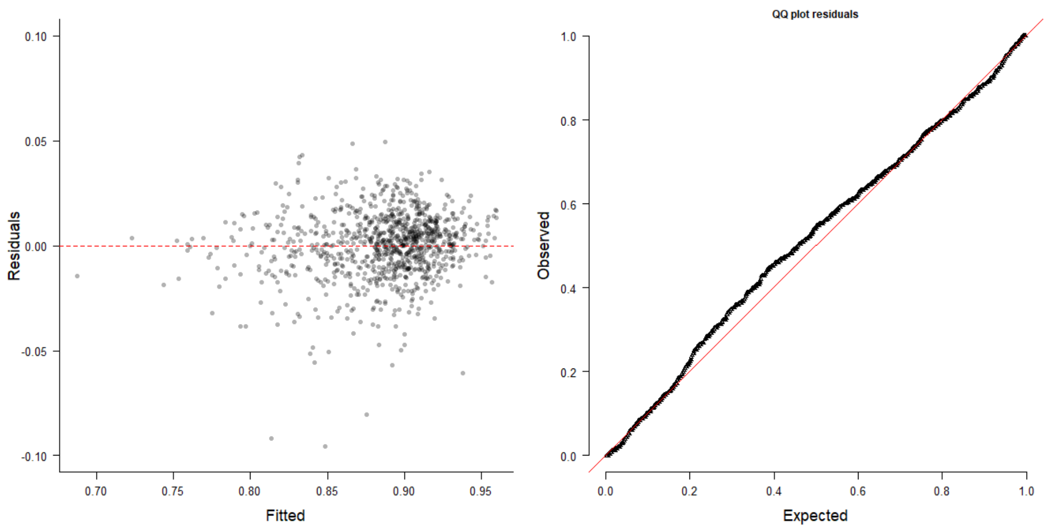


*Figure S5: Diagnosis of the regression on the vaccine coverage. Left panel: Fitted vs residuals plot, Left panel: uniform quantile-quantile plot.*

Finally, the stratification of residuals by year did not show any trend, which indicates the fit was consistent throughout the different years included in the dataset (Figure S6).

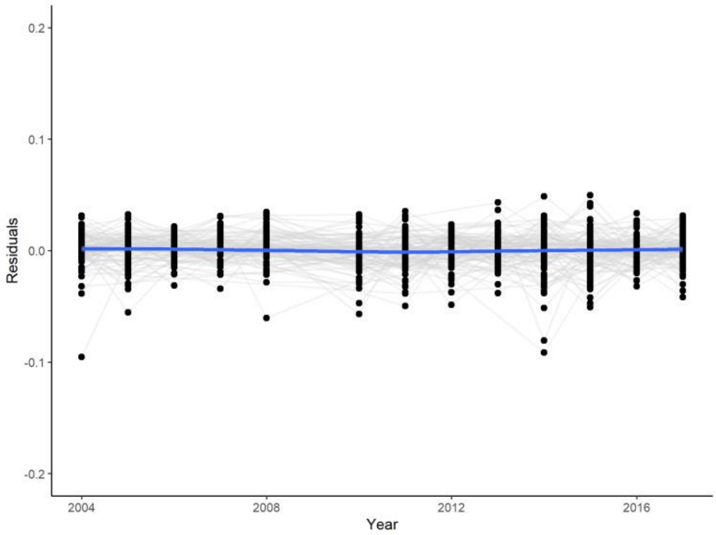


*Figure S6: Distribution of the residuals per year of inference. The blue line indicates the mean value every year.*

The multiple diagnostics tests and plots we used mostly supported the specification of the beta regression model. We inferred the missing values of coverage by using the mean estimates of the model for the years and regions where data was missing. Since the diagnostics also indicated minor discrepancies between our model and the results, we also generated 100 sets of coverage data by drawing the missing data from the normal distribution of the model (using the mean and standard deviation of the inferred values for the missing entries). We then ran the hhh4 models on each of the full coverage datasets to highlight the influence of the missing entries on parameter estimates. The deviation from the parameter estimates generated with the mean coverage was minimal (Figure S7 and

Figure S8). This indicates that our conclusions are robust to changes in the inferred values of the missing data.

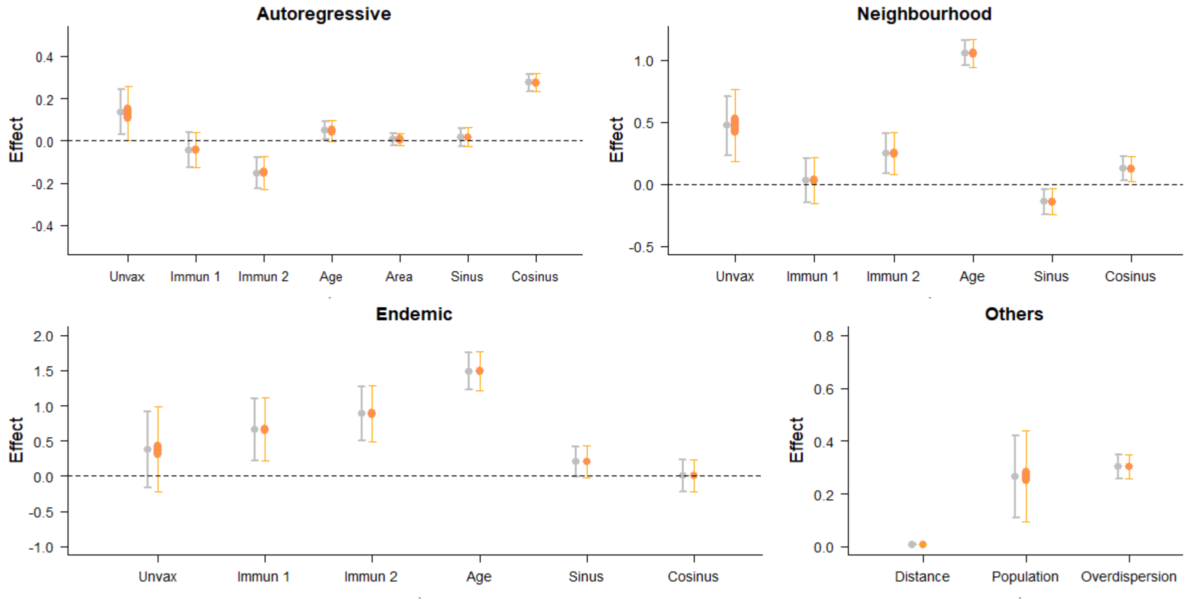


*Figure S7: Estimates of the parameters in each component of Model 1, using the reference fit (grey) and 100 different values of coverage for the inferred entries (orange). unvax corresponds to the effect of $u_{i,t}$, the mean proportion unvaccinated over the three years before t in i; incid 1 and incid2 correspond to the effect of $N^1_{i,t}$ and $N^2_{i,t}$ the category of incidence in the three years before t in i; pop corresponds to the effect of $m_{i,t}$ the number of inhabitants at t in i; area corresponds to the effect of the surface; sin and cos correspond to the effects of seasonality; distance and population correspond to the spatial parameters of the connectivity matrix w (δ and γ); overdisp is the estimate of the log-overdispersion parameter in the negative binomial distribution of $Y_{i,t}$. Dots show the mean values associated with the parameters; arrows show the 95% Confidence interval. The orange arrows indicate the extreme values of the 95% confidence interval obtained drawing different values of coverage for the missing entries.*

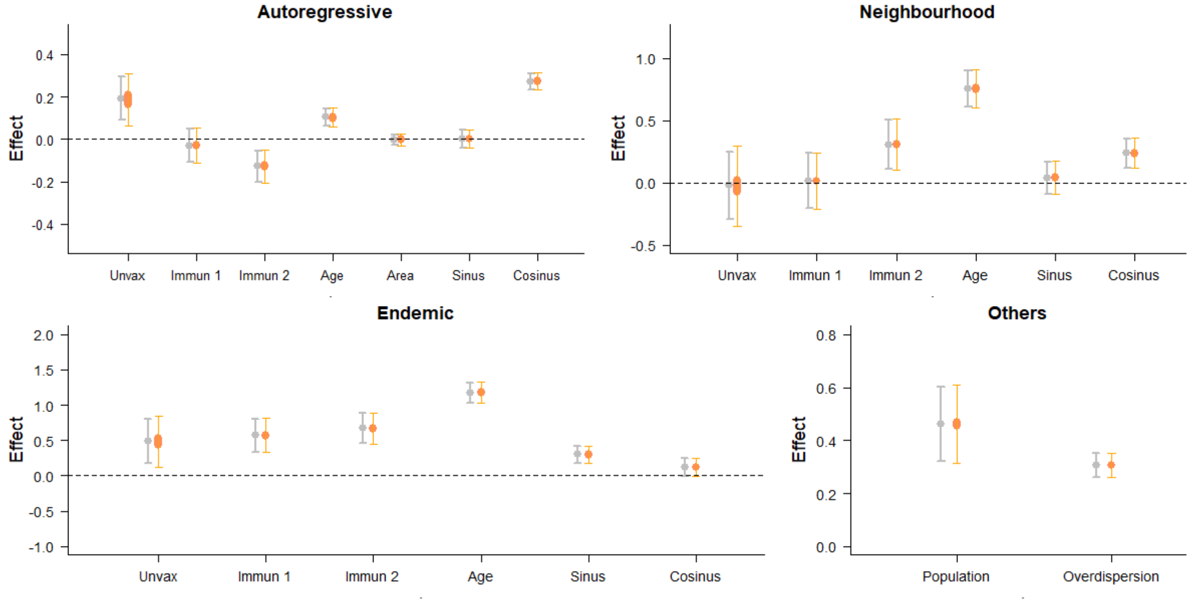


*Figure S8: Estimates of the parameters in each component of Model 2, using the reference fit (grey) and 100 different values of coverage for the inferred entries (orange). unvax corresponds to the effect of $u_{i,t}$, the mean proportion unvaccinated over*

*the three years before t in i; incid1 and incid2 correspond to the effect of $N_{i,t}^1$ and $N_{i,t}^2$ the category of incidence in the three years before t in i; pop corresponds to the effect of $m_{i,t}$ the number of inhabitants at t in i; area corresponds to the effect of the surface; sin and cos correspond to the effects of seasonality; distance and population correspond to the spatial parameters of the connectivity matrix w (δ and γ in Equation X); overdisp is the estimate of the log-overdispersion parameter in the negative binomial distribution of $Y_{i,t}$. Dots show the mean values associated with the parameters; arrows show the 95% Confidence interval. The orange arrows indicate the extreme values of the 95% confidence interval obtained drawing different values of coverage for the missing entries.*

### 3. Seasonality

Both Model 1 and Model 2 include two parameters per component describing the seasonality of transmission and importations. For each component and each model, we computed $\exp\left(\beta_c^{(\lambda)} cos\left(\frac{2\pi t}{365}\right) + \beta_s^{(\lambda)} sin\left(\frac{2\pi t}{365}\right)\right) - 1$, the multiplicative factor corresponding to the impact of seasonality on the predictor (Figure S9).

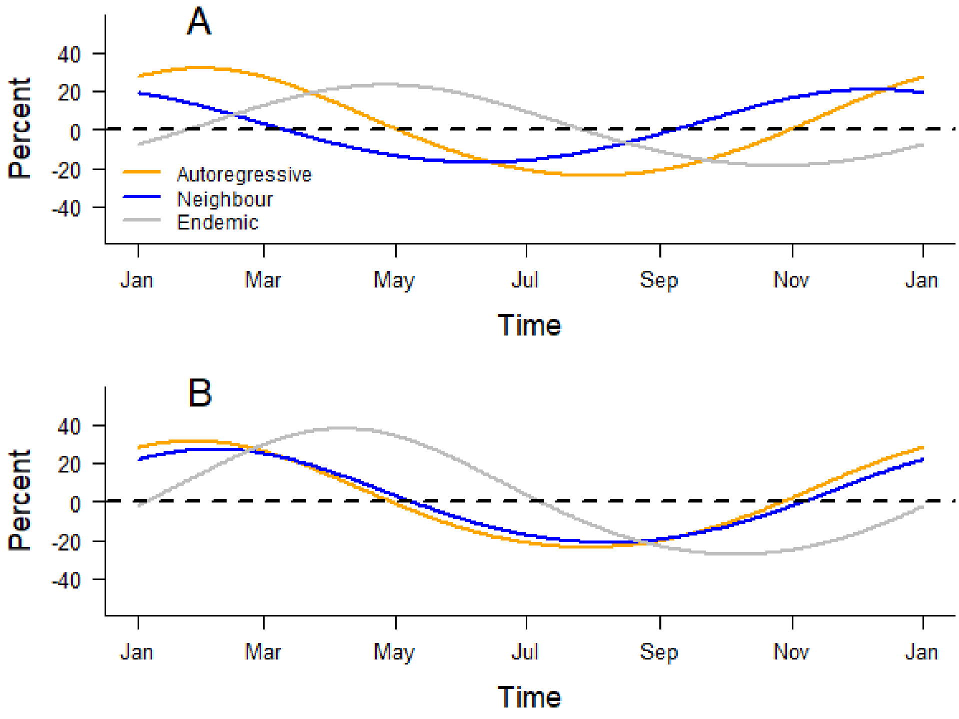


*Figure S9:Seasonality of each component in Model 1 (Panel A) and Model 2 (Panel B). We quantified the impact of seasonality using the percent of variation around the mean value every day.*

### 4. Analysis using the neighbour-based connectivity matrix

The calibration and simulation study presented in the Main text was also run using Model 2. The fits to daily and weekly data were similar to Model 1 (Figure 4 and Figure S10). The calibration study indicates that Model 2 was slightly more likely to underestimate the number of cases in short-term predictions than Model 1. We generated the national number of cases predicted 3, 7, 10, and 14 days ahead by Model 1 and Model 2 over the calibration period, and compared the forecasts to the data (Figure S11). The predictions in both models were very similar, with the 95% prediction intervals overlapping on the majority of the calibration period. The data points were included in the 95% prediction intervals for forecasts one week ahead or less, when the period of forecasts was 10 or 14 days, we observed a lag between the predictions and the data when the number of cases started increasing and dropping.

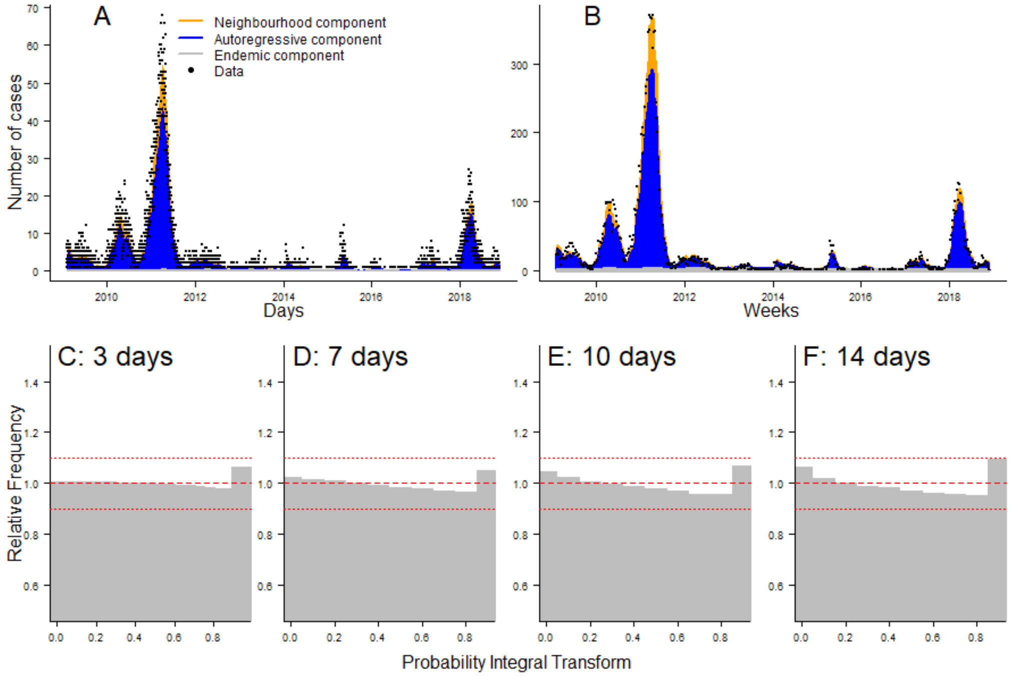


*Figure S10: Panel A and B: Daily and weekly fit between the data and Model 2. The inferred number of cases is split among the three components of the model. Panel C to F: PIT histograms of Model 2, generated respectively for predictions 3, 7, 10, and 14 days ahead.*

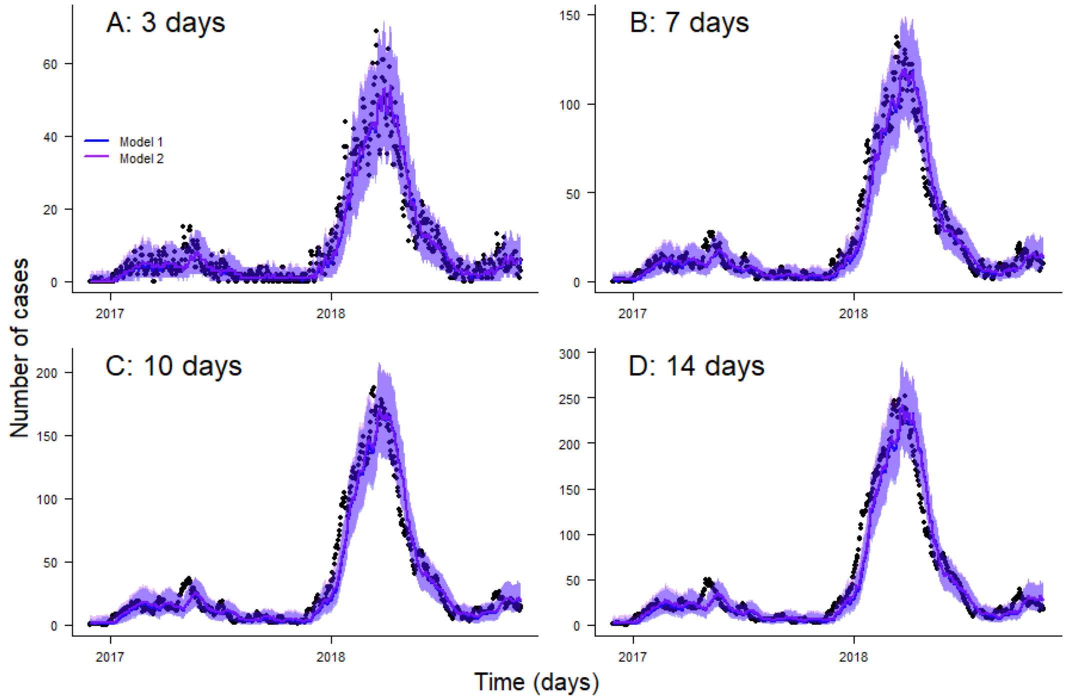


*Figure S11: Comparison between predictions 3, 7, 10, and 14 days ahead using model 1 and model 2 and the data. Lines correspond to the median estimates, shaded areas correspond to the 95% predictions intervals. The blue and purple predictions are similar for the entire calibration period, hence the curves overlap in all panels. Black dots represent the number of cases 3, 7, 10, and 14 days ahead in France at each date.*

The proportion of cases that stem from the endemic component was slightly higher in Model 2, which is due to the framing of the neighbourhood component (Figure S12). Indeed, in Model 2, the neighbourhood component only contains transmission between neighbours, whereas in Model 1, it describes any cross-regional transmission between the regions included in the study. Therefore, long-distance transmissions fall into the endemic component in Model 2, whereas they would be included in the neighbourhood component of Model 1.

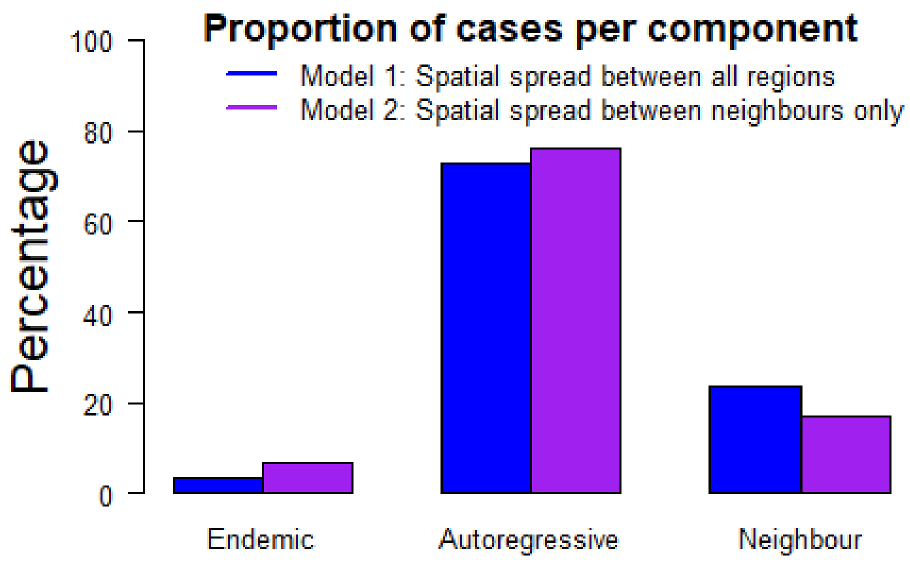


*Figure S12: Proportion of cases per component in both models*

We generated four simulation sets using the scenarios presented in the Main text with the parameter estimates from Model 2. We observed similar spatial heterogeneity in exposure and risk of large outbreaks. The areas most likely to be affected by large outbreaks were Paris and its suburbs, along with the south of France (Figure S13). Setting the level of recent incidence to the minimum values in each region decrease the number of baseline importations, and therefore reduced the proportion of simulation where most regions were exposed to transmission (i.e. they reported at least one case). Nevertheless, the risks of large outbreaks were similar to the reference simulation set. The effect of variations in vaccine coverage on the risks of importations and local transmission was the same as what was described for Model 1, with a three percent decrease leading to an abrupt increase in the number of cases.

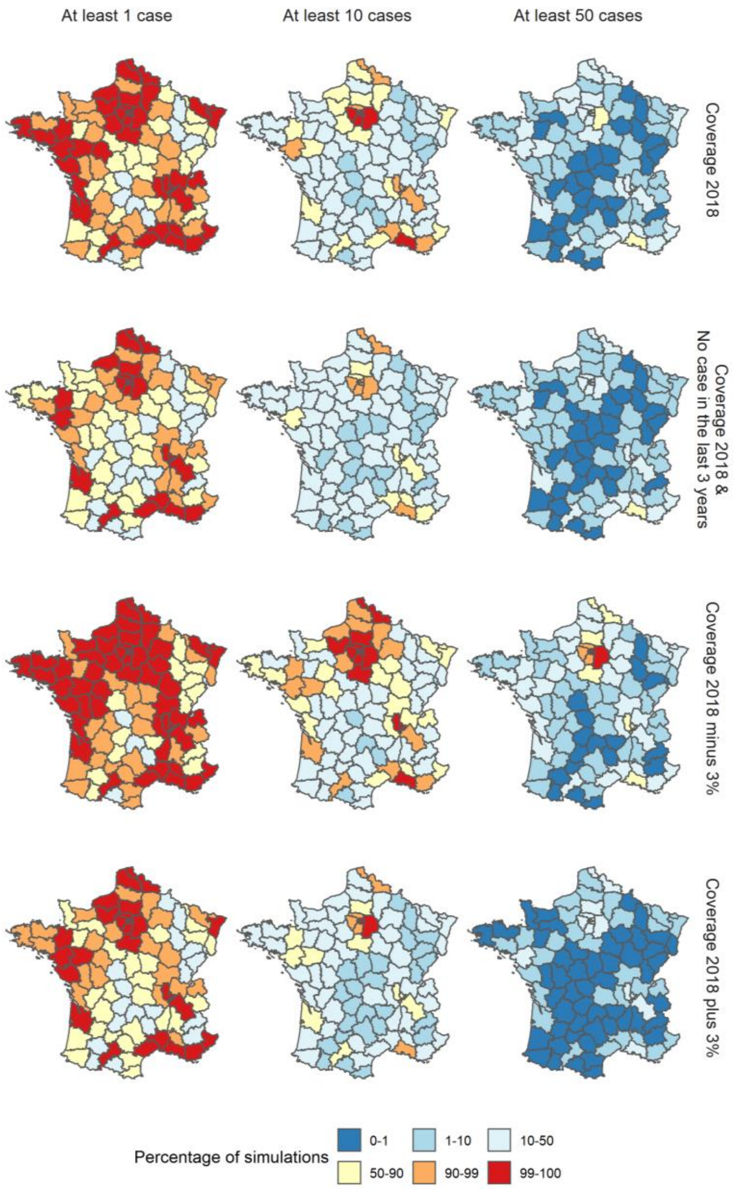


*Figure S13: Percentage of simulations where the number of cases reported in each region in 2019 was at least 1, 10, and 50 cases for each scenario using parameter estimates from Model 2. Each row corresponds to a different scenario: i) Reference, ii) Minimum level of recent incidence in each region, iii) Local vaccine coverage increased by 3% in each region, iv) Local vaccine coverage decreased by 3% in each region.*

The spatial spread upon repeated importation was more limited than in Model 1 (Figure S14). In all four scenarios, most regions were not exposed to transmission in any of the simulations. This is due to the fact that long-distance transmission would fall into the endemic component of Model 2, which was not used for these simulation sets. On the other hand, large transmission clusters in the region of importations and its neighbours were more common.

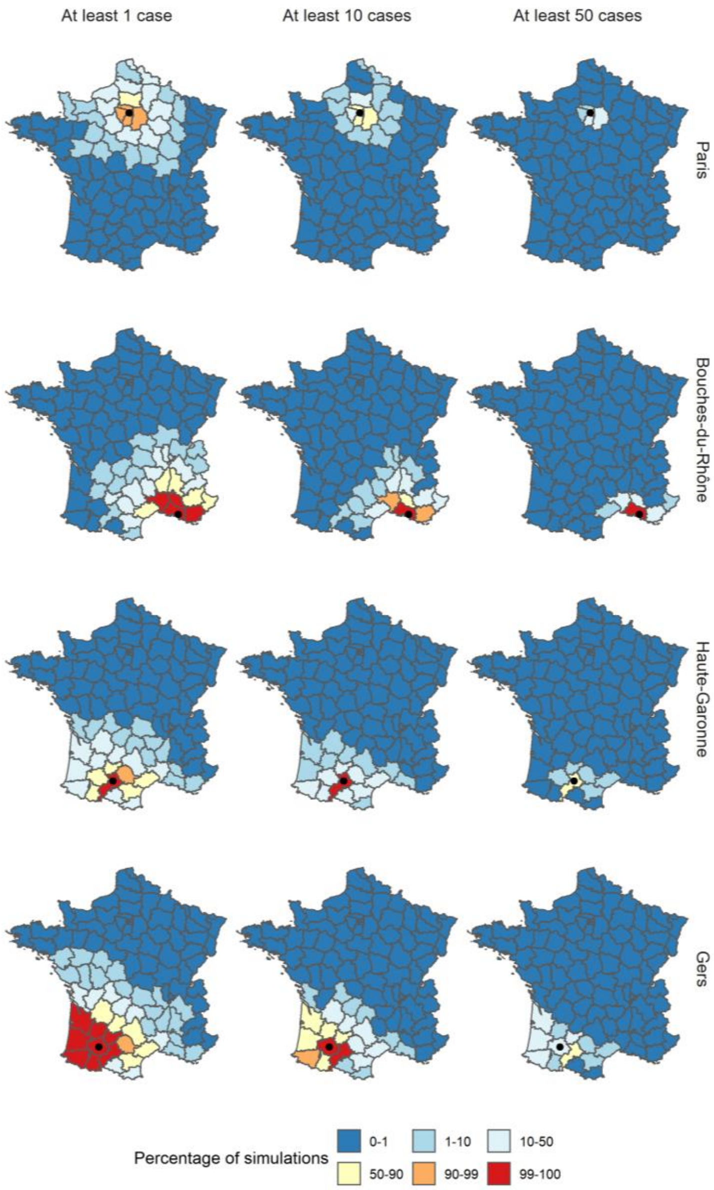


*Figure S14: Percentage of simulations where the number of cases reported in each region in 2019 was at least 1, 10, and 50 cases following the importations of ten cases in December 2018, and using the parameter estimates from Model 2. For each row, the region of importation is indicated by a black dot.*

### 5. Last values of the covariates

The simulation sets were generated using the last measures of the variables included in our models. We show the geographic distributions of each variable (Figure S15):

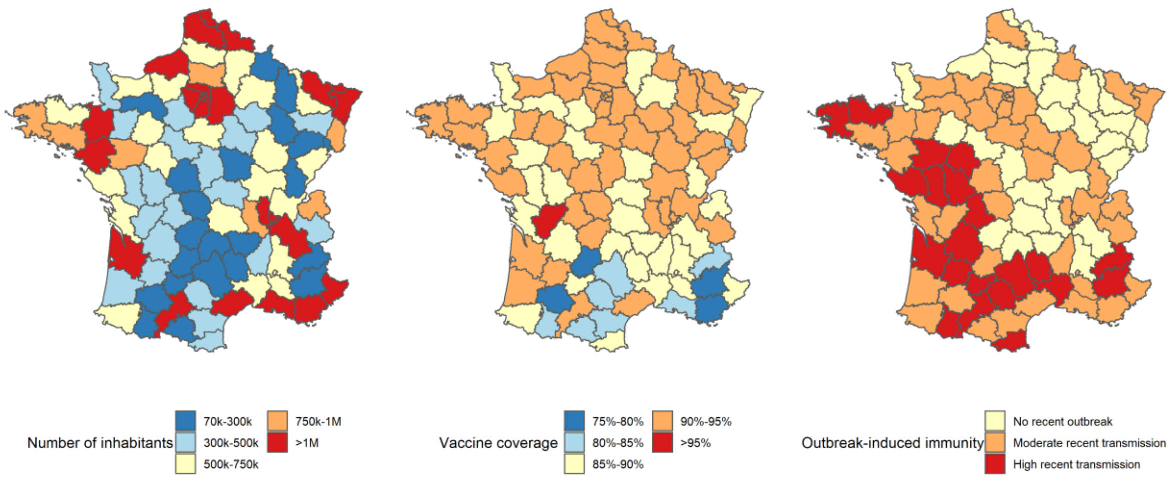


*Figure S15: Geographic distribution of the number of inhabitants (right panel), average vaccine coverage (central panel), and recent incidence (left panel) at the end of 2018.*

### 6. Local importations with different vaccine coverage

Decreasing the vaccine uptake in each region by three percent led to an increase in the number of regions exposed to transmission following a local group importation (Figure S16). The risks of generating large outbreaks were higher both in the region of imports and in other areas, especially in highly populated urban regions. This was due to an increase in the number of cases generated in the neighbouring component, which led to more cross-transmission into regions with higher number of inhabitants.

On the other hand, a reduction of vaccine uptake led to a decrease in the overall number of cases generated per outbreak (Figure S17). Spill overs from the region of origin were much rarer: in the case of group importations in Haute-Garonne or Bouches-du-Rhône, no department outside the department of importation was exposed to one case in more than half of the simulations. Risks of large outbreaks were also reduced: in all four simulations, no department reported more than 50 cases in at least half the simulations, and few apart from the department of importation reported more than 10 cases.

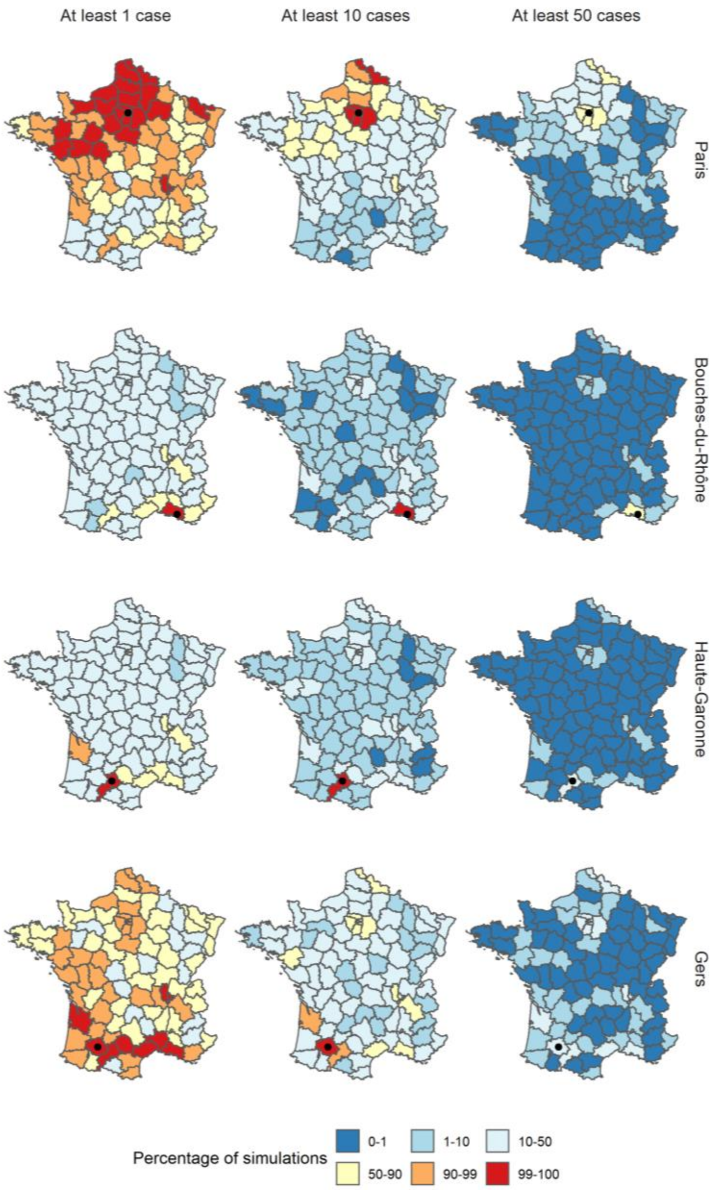


*Figure S16: Percentage of simulations where the number of cases reported in each region in 2019 was at least 1, 10, and 50 cases following the importations of ten cases in December 2018, and using the parameter estimates from Model 1 and a three percent decrease in vaccine coverage in each region. For each row, the region of importation is indicated by a black dot.*

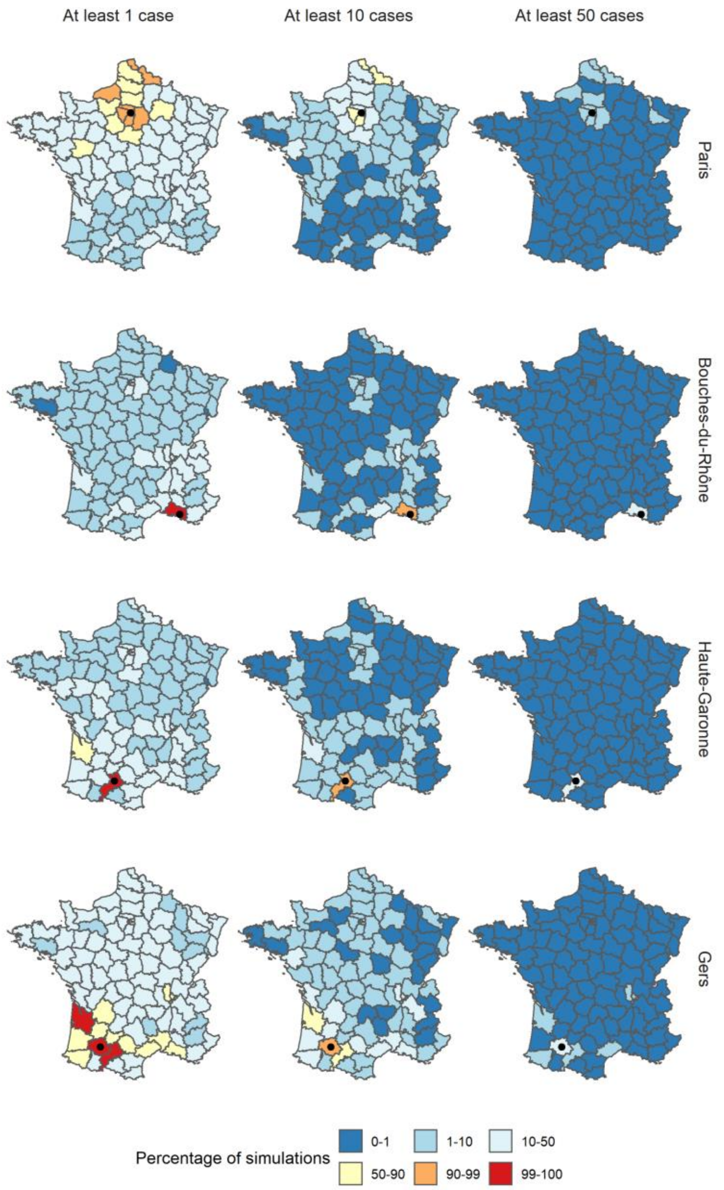


*Figure S17: Percentage of simulations where the number of cases reported in each region in 2019 was at least 1, 10, and 50 cases following the importations of ten cases in December 2018, and using the parameter estimates from Model 1 and a three percent increase in vaccine coverage in each region. For each row, the region of importation is indicated by a black dot.*

### 7. Comparison with aggregated models and impact random effects

We assessed the impact of using daily case count and random effects on the calibration of the models by the Ranked Probability score. We were interested in three models: The daily model without random effects (presented in the Main Text), the daily models with random effects, and the aggregated model, fitted using a 10-day aggregation. Although random effects allow for more flexibility in the model, they significantly slow down the fitting procedure (40 times slower [2]). All models were run with the same specifications as Model 1, where cross-regional transmission can happen between every region, and with using vaccine coverage, recent levels of transmission, number of inhabitants, surface, and seasonality as covariates.

For each of the three models, we generated 10-day predictions every 10 days for the last two years of data. This corresponded to 72 calibration dates for 94 regions, hence 6,768 data points. We computed three different indicators using the R package scoringutils [3,4]:

1. The sharpness shows the ability of the model to generate predictions in a narrow range of possible outcomes, which means that the sharpness score is independent of the data. We used the normalised median absolute deviation about the median.
2. The bias: indicates whether a model systematically under or over predicts. Least biased models will get a mean value around 0, whereas completely biased models will get a value of -1 or 1.
3. The average Ranked Probability Score (RPS) for Count Data, proper scoring rule minimised if the predictive distribution is the same as the one generating the data.

The daily model without random effect had the lowest value of sharpness, indicating that the forecasts were generated in a narrower range of outcomes than the other models (Figure S18). The mean value of bias was closest to 0 in the aggregated model, which show that these forecasts were well balanced and did not tend to under or over-estimate the number of cases. We performed a permutation test on the RPS score, which indicated that the calibration of the two daily models was significantly better than the aggregated model (p-value < 0.02) [2].

Adding the random effects to the daily model did not lead to major improvements: The sharpness and bias were better in the model without random effects, whereas the difference between the RPS scores was not significant (p-values > 0.1). Since using random effects slows down the fitting procedure, without contributing a major improvement to the calibration, random effects were not integrated into the main analysis.

The fact that random effects did not substantially improve the calibration may be due to the time span of this analysis: random effects can be used to account for heterogeneity between the regions that would not be explained by the covariates included in the analysis. Since a random effect is applied to a given region throughout the entire time span of the data, they quantify constant effects on the region that was otherwise unobserved. As this analysis covers almost ten years of data, the impact of factors not included in this analysis may have varied over time, which would explain the little added value of the random effects.

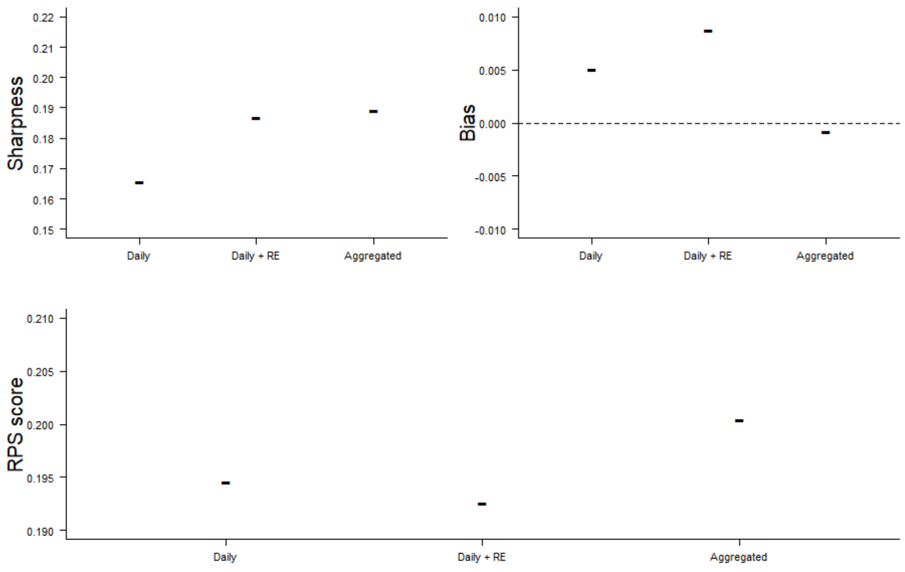


*Figure S18: Sharpness, bias and RPS scores of the Daily model with and without Random Effects (RE), and of the aggregated model*

### 8. Control for day-of-the-week effect

Since we use daily onset dates, we explored the impact of potential reporting bias based on the day of the week. Indeed, delays in reporting can cause bias in the date of declaration of cases, and can explain why more cases would be reported on weekdays than on weekends. The number of cases with an onset date on Saturdays or Sundays was slightly lower than for the other days (1,970 cases on Saturdays and Sundays, whereas the average value on weekday is 2,100 cases, Figure S19).

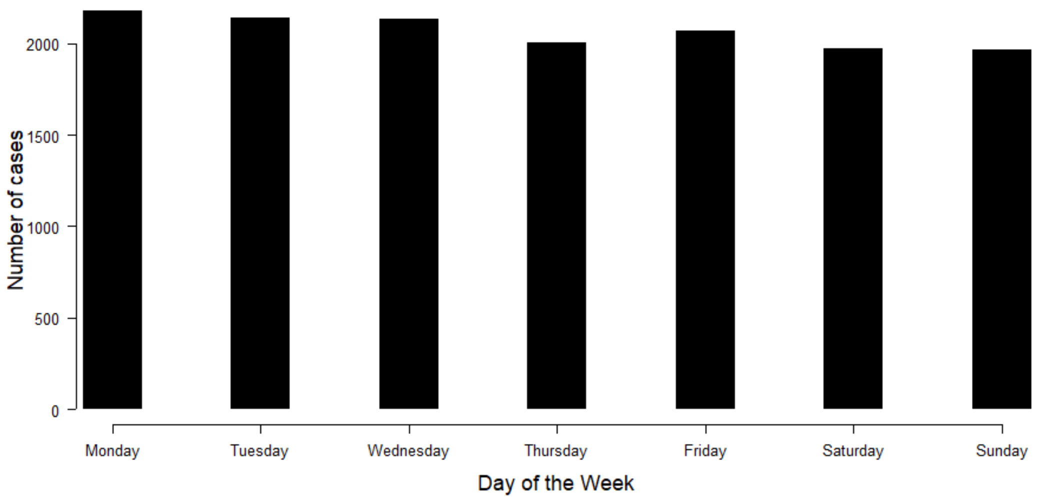


*Figure S19: Number of cases by day of the week. We used the onset date for each case reported to the ECDC between 2009 and 2018.*

We implemented a model controlling for the weekend effect in each component. This model contained the same covariate and distance matrix as Model 1. The covariate "Weekday" was a binary variable, equal to 1 on Saturdays and Sundays, and 0 otherwise. In the neighbourhood and endemic components, the covariate "weekday" had no significant impact on the value of the predictor (Figure S20). On the other hand, the number of cases stemming from the autoregressive component was smaller on weekends (coefficient estimate: -0.09 [-0.15- -0.03]). Introducing this covariate brought no change to the values of the other coefficients in the model.

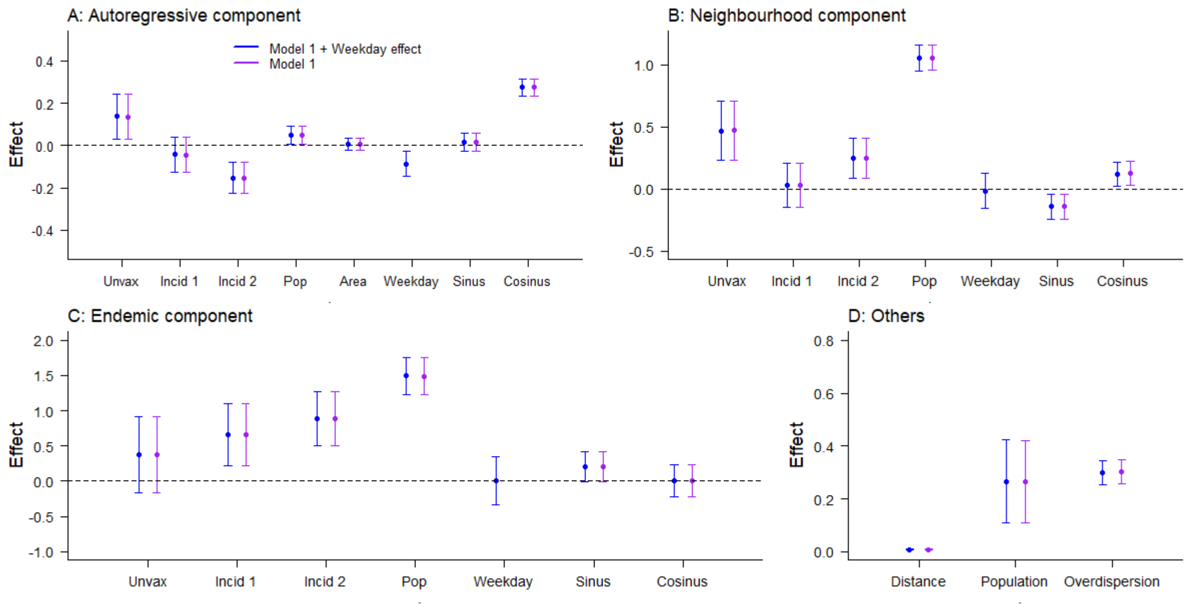


*Figure S20: Comparison of the parameter estimates obtained in Model 1 (similar to Figure 2) with or without a weekday covariate added in each compartment. Weekday was computed as a binary covariate, whose value was 1 on Saturdays and Sundays, and 0 otherwise.*

### Reference


[1] Brooks ME, Kristensen K, van Benthem KJ, Magnusson A, Berg CW, Nielsen A, et al. glmmTMB balances speed and flexibility among packages for zero-inflated generalized linear mixed modeling. R J 2017;9:378–400. doi:10.32614/rj-2017-066.

[2] Meyer S, Held L, Höhle M. hhh4: Endemic-epidemic modeling of areal count time series. J Stat Softw 2016.

[3] Funk S, Camacho A, Kucharski AJ, Lowe R, Eggo RM, Edmunds WJ. Assessing the performance of real-time epidemic forecasts: A case study of Ebola in the Western Area Region of Sierra Leone, 2014–15. BioRxiv 2017:1–17. doi:10.1101/177451.

[4] Bosse NI, Abbott S, EpiForecasts, Funk S. scoringutils: Utilities for Scoring and Assessing Predictions 2020. doi:10.5281/zenodo.4618017.
